# Artificial intelligence for pulmonary tuberculosis detection on chest X-ray images: A systematic review and diagnostic accuracy meta-analysis

**DOI:** 10.64898/2026.11.11.26367872

**Authors:** Sayem Al Mahfuz, Shimanto Kumar Roy, Hamim Ibne Rahman, Aminun Nahar

**Affiliations:** Cox’s Bazar Medical College, Cox’ Bazar, Bangladesh; BRAC University, Dhaka, Bangladesh; American International University Bangladesh, Dhaka, Bangladesh

## Abstract

Artificial intelligence (AI)-based computer-aided detection (CAD) of tuberculosis (TB) on chest X-ray (CXR) may standardize screening and triage in settings with few radiologists but its accuracy in clinical cohorts remains uncertain. We searched five databases (PubMed, ScienceDirect, IEEE Xplore, OpenAlex, Lens.org) on 15 March 2026 for studies published between January 2020 and March 2026, and we appraised these studies using QUADAS-2. Evaluations from clinical cohorts with complete 2 × 2 data and a microbiological or composite reference standard for all participants were pooled using a bivariate random-effects model, with 95% prediction intervals. We accounted for overlapping participants by performing a post hoc analysis with one evaluation per cohort and used GRADE to assess the certainty of the evidence. No protocol was prepared or registered. We included 21 studies (26 evaluations of AI models). Nine evaluations from three independent cohorts were pooled.

Pooled sensitivity was 0.896 (95% confidence interval [CI] 0.874 to 0.915, prediction interval 0.798 to 0.950) and pooled specificity was 0.689 (95% CI 0.642 to 0.732; prediction interval 0.499 to 0.831). Both confidence intervals include the World Health Organization (WHO) triage thresholds (sensitivity 90%, specificity 70%). When a screening study with partial verification was added, pooled specificity increased to 0.735. Eighteen of 24 assessed evaluations were at high risk of bias and only 1 of 21 studies was prospective, fully verified, externally validated and met the WHO thresholds. Certainty of evidence was low for sensitivity and very low for specificity. A subsequent PubMed search found additional eligible clinical evaluations that were not screened, so the review is incomplete for those studies. In clinical cohorts, AI-CXR sensitivity is close to the WHO triage target, and specificity is close to, but not clearly above, its target. The evidence is pooled across only three cohorts, and specificity varies by product and setting. Thresholds need to be calibrated locally, and prospective, externally validated evaluations are needed before widespread deployment.

## 1. Introduction

### 1.1 Global burden of tuberculosis

Tuberculosis (TB) is preventable, treatable but it is still the leading infectious killer in the world. The World Health Organization (WHO) estimates that 10.7 million people fell ill with TB in 2024 (5.8 million men, 3.7 million women and 1.2 million children aged 0–14 years) and that 1.23 million people died of the disease [1].

*Mycobacterium tuberculosis* transmission, delayed diagnosis, limited access to confirmatory testing, under-recognition of smear-negative disease and limited capacity for systematic screening in high-burden settings all contribute to the persistent burden [1, 2]. The populations examined in the evidence reviewed here (symptomatic adults, community-screening participants, emergency-department patients and smaller pediatric cohorts) are precisely those in whom missed or delayed diagnosis carries major consequences [3–9].

Diagnosis is especially challenging in settings where microbiological testing or radiological expertise is not available. Studies were sourced from Pakistan, Bangladesh, China, the Philippines, Taiwan, India, Mozambique, Spain and elsewhere, revealing a growing but uneven global evidence base on artificial intelligence (AI)-assisted chest X-ray (CXR) interpretation [3–8, 10]. Children, people living with HIV, those with smear-negative disease, older adults and those with previous TB require special attention because their radiographic and microbiological presentations are different from those of adults with smear-positive pulmonary TB. The evidence presented supports this concern: sensitivity decreased for smear-negative TB, specificity was lower in older adults and in those with previous TB in evaluations of commercial computer-aided detection (CAD) software, and the only dedicated pediatric study showed a clear decline on external validation [3–5, 7, 8].

### 1.2 Chest X-ray for TB screening

CXR is fast, widely available and can detect radiographic abnormalities before or alongside microbiological confirmation [2]. WHO screening guidance recognizes CXR and CAD as tools that identify individuals who need confirmatory testing with WHO-recommended rapid molecular diagnostics [2, 11]. In a large evaluation in Bangladesh, the use of AI interpretation of CXR as a triage step before Xpert MTB/RIF testing reduced the number of Xpert tests required while maintaining sensitivity close to the WHO triage target [4].

However, interpretation of CXR is influenced by the training of the reader, image quality, disease spectrum and the overlap of TB radiographic findings with healed TB, other infections and chronic lung disease. Thus, the studies included showed a large variation in specificity, especially in real-world case finding and among people with previous TB, older age or low body mass index [3, 4, 7].

### 1.3 Artificial intelligence for interpretation of CXR

Deep-learning systems learn visual patterns from large collections of images, and then output a probability or abnormality score for TB-compatible findings. The literature reviewed here includes commercial CAD systems, Custom convolutional Neural Networks (CNNs), transformer-based and hybrid networks, Graph Neural Networks (GNNs), segmentation-classification pipelines and explainable AI (XAI) approaches [4, 7, 8, 10, 12–24].

Commercial tools (qXR, CAD4TB, Lunit INSIGHT CXR, InferRead DR and JF CXR-1) are the most relevant for implementation because they have been evaluated in clinical or screening cohorts rather than only on public benchmarks. In Pakistan, Khan et al. prospectively evaluated qXR and CAD4TB against culture [3]. Qin et al. compared five products in a large Bangladeshi triage cohort [4]. Liao et al. evaluated JF CXR-1 in active screening in China [5]. Tavaziva et al. evaluated Lunit INSIGHT against culture [7]. Marquez et al. evaluated qXR in Philippine case finding [6]. In parallel, research models (NFNet, custom CNNs, multi-stage segmentation and feature-fusion pipelines, CapsNet–SOM, SPX-GNN, EfficientNet–ViT hybrids and multi-scale CNNs) often demonstrate very high accuracy on curated public datasets whose class balance, image quality and labels may not represent clinical screening [10, 14, 16, 18, 20, 22–24]. Explainability methods such as Grad-CAM, LIME, SHAP and perturbation-based importance maps have been used to show which image regions drive model output [12, 13, 23–25].

### 1.4 Evidence gaps at present

The evidence base is heterogeneous regarding populations, datasets, operating thresholds, model types and reference standards. Reference standards include culture, Xpert MTB/RIF, composite clinical definitions and dataset labels, so apparent differences in performance may reflect study design rather than model quality [3–7, 10, 17]. External validation is a major weakness. A model trained on the Shenzhen dataset performed substantially worse on an external NIH population, and pediatric performance declined on an external low-burden cohort [8, 17]. Many experimental studies used public or Kaggle datasets with artificial class balance and limited demographic information [14, 16, 18, 21–24]. Reporting is not standardized. Predictive values, calibration, precision–recall curves and complete 2 × 2 tables are frequently missing, although they determine referral burden in screening programs.

Children and people with HIV are underrepresented, and explanations are rarely validated against radiologist judgment [12, 13, 23, 25].

### 1.5 Objectives and research questions

The main aim of this review, reported following PRISMA 2020 [26], was to assess the diagnostic accuracy of AI-based systems for detecting pulmonary TB on CXR in studies published from 2020 to 2026. Secondary objectives were to (i) compare model families (ii) appraise risk of bias and applicability with QUADAS-2 [27] (iii) quantify the effect of dependence between evaluations drawn from the same cohort on pooled estimates and (iv) characterize the evidence needed for clinical translation.

This review asked six questions:

**RQ1.** What is the reported diagnostic performance of AI-based detection of pulmonary TB from CXR (2020–2026)?

**RQ2**. What are the dominant model families, datasets, populations, validation strategies and reference standards in the evidence base?

**RQ3**. How consistently are clinically important metrics (sensitivity, specificity, area under the receiver operating characteristic curve [AUC], predictive values, calibration, 2 × 2 counts) reported?

**RQ4**. What are the main sources of bias and applicability concern?

**RQ5.** How do commercial CAD systems differ from research models in accuracy and in evidence for translation?

**RQ6**. What methodological and clinical gaps must be addressed before AI-CXR can be safely incorporated into TB screening and triage?

## 2. Background

### 2.1 Standard TB screening and diagnosis

The WHO End TB Strategy targets continue to be difficult to reach in high-burden settings [1, 28]. The standard sequence involves symptom screening, CXR and, for those with an abnormal result, microbiological confirmation by Xpert MTB/RIF or another WHO-recommended rapid molecular test [2]. CXR is cheap and rapid, can detect abnormalities before microbiological confirmation [29] and is recommended by WHO for triage [30]. Its main limitation is the need for a trained reader. The density of radiologists is very low in many low-and middle-income countries, which motivates automated interpretation that can be operated by minimally trained staff.

### 2.2 AI methods for medical image analysis

Convolutional neural networks use layered filters that progressively detect edges, textures and shapes, allowing recognition of opacities, cavitation and other TB-compatible findings.

Vision transformers model the relationships between patches of an image with self-attention to capture broader spatial context. Transfer learning takes networks trained on large generic datasets (e.g. ImageNet) and adapts them to medical tasks, which is useful when labeled data are scarce. Convolution–transformer hybrids (e.g. CoAtNet) combine convolutions for feature extraction with transformers for attention; in this review they are grouped with transformer-based models (Section 4.2.5). Capsule networks and graph neural networks preserve spatial relationships between features or model images as graphs of connected regions, capturing long-range dependencies.

### 2.3 Deep-learning architectures for TB detection

Bhandari et al. developed a deep-learning model with explainable AI that classified CXRs into four classes (COVID-19, pneumonia, TB and normal) with an average test accuracy of 94.3% [12] Acharya et al. achieved 96.9% accuracy and an AUC of 0.994 with their NFNet [10]. Wang et al. used an EfficientNetV2 ensemble in an emergency-department population [25]. Siddharth et al. achieved 86.4% accuracy and an AUC of 0.938 with CoAtNet on an Indian national survey dataset [13]. Capellán-Martín et al. developed pTBLightNet, a multi-view pediatric detector that uses frontal and lateral CXRs [8]. Pala et al. modeled images as superpixel graphs and reached almost perfect accuracy on a benchmark dataset [23]. Sharmila et al. combined capsule networks with self-organizing maps (93.6% accuracy; the authors frame the task as multidrug-resistant TB detection, but the public datasets used carry TB-versus-normal labels) [15], and Maashi et al. proposed a multi-stage segmentation and feature-fusion framework (98.3% accuracy on a 600-image dataset) [14].

### 2.4 Commercial CAD systems for TB screening

Unlike research prototypes, commercial CAD systems are built for daily use. qXR (Qure.ai) is among the most evaluated products; in Philippine case finding it achieved high apparent sensitivity but very low apparent specificity at a fixed threshold [6]. CAD4TB (Delft Imaging) provides a continuous score, and the threshold is left to the user. At a common operating point of 90% sensitivity, Lunit INSIGHT CXR had a specificity below 70% [4], and in a culture-referenced cohort it had a sensitivity of 88.1% and a specificity of 57.9% [7]. JF CXR-1 (JF Healthcare) and InferRead DR (Infervision) were less specific than qXR and CAD4TB at matched sensitivity [4]. In a low-prevalence screening program, JF CXR-1 identified all eight TB cases at a 30% threshold with 95.7% specificity, although not all participants underwent microbiological testing [5]. Commercial products are trained on proprietary data, their performance varies with population, equipment, prevalence and threshold, and few have been evaluated prospectively.

### 2.5 Explainable AI for medical imaging

Grad-CAM produces heatmaps of the regions that influence a prediction; SHAP quantifies the contribution of features; LIME perturbs regions of the image to estimate their local importance; and perturbation-based methods infer importance by systematically altering regions. Five of the included studies used XAI [12, 13, 23–25], and only one validated the explanations with radiologists [12]. Without such validation, heatmaps are illustrative, and AI output remains decision support, not diagnosis.

### 2.6 Existing reviews and open gaps

An early review highlighted the absence of clinical evaluation, the dependence on case– control designs and weak reference standards [31]. Recent reviews pooled the sensitivity and specificity of certified products and reported estimates approaching WHO triage targets with substantial heterogeneity [32, 33]. Two additional syntheses are directly relevant. A systematic literature review of machine-and deep-learning TB detection on CXR, with a meta-analysis of ten studies [34], and an individual-patient-data meta-analysis of commercial deep-learning software used as a triage test [35]. Important gaps remain: incomplete metric reporting, inconsistent application of QUADAS-2, limited synthesis of performance in active screening, emergency, pediatric and HIV-affected populations, little appraisal of XAI validation, variable operating thresholds and, in quantitative syntheses, frequent treatment of multiple algorithms evaluated on the same participants as independent studies. This review addresses these gaps with a synthesis reported according to PRISMA 2020 that distinguishes commercial from research models, explicitly deals with cohort-level dependence and reports prediction intervals in addition to confidence intervals.

## 3. Methods

### 3.1 Reporting guidelines, protocol and post hoc changes

This review was reported in accordance with PRISMA 2020 and the diagnostic test accuracy extension PRISMA-DTA [26, 36]. The completed PRISMA 2020 checklist is provided as S1 Checklist and the PRISMA-DTA checklist as S2 Checklist. No protocol was prepared in advance, and none was registered. The review question, eligibility criteria, sources of information, search strategy, screening workflow, extraction variables, QUADAS-2 process and synthesis plan were specified during the conduct of the review. This makes it impossible to verify from the outside that the analyses were specified before looking at the data, and it increases the risk of selective reporting. We therefore list every change made after data inspection, including those made in response to peer review, in Table 1, and we label such analyses as post hoc throughout. Ethics approval and consent were not required because only published aggregate data were used.

**Table 1.**
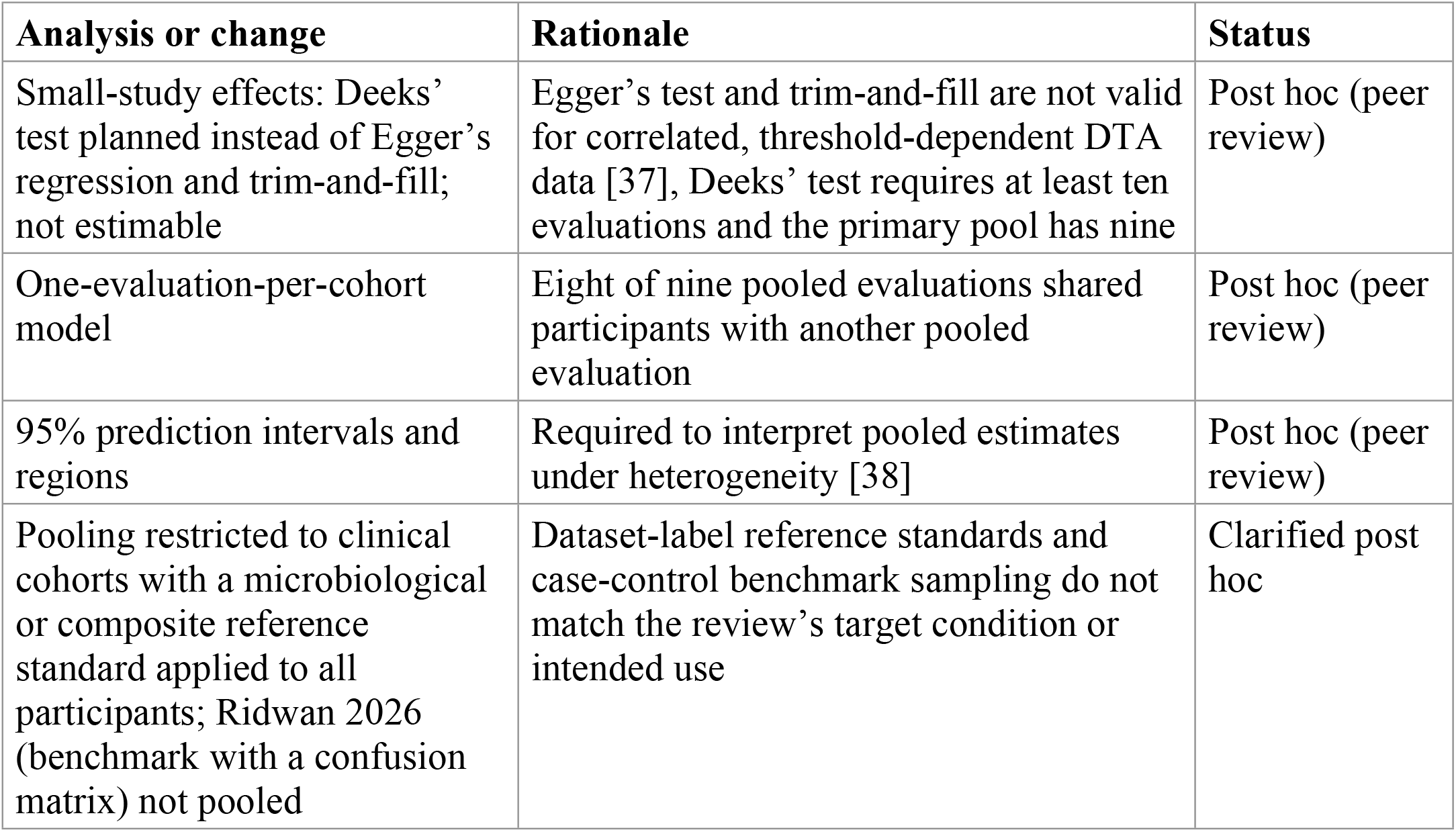
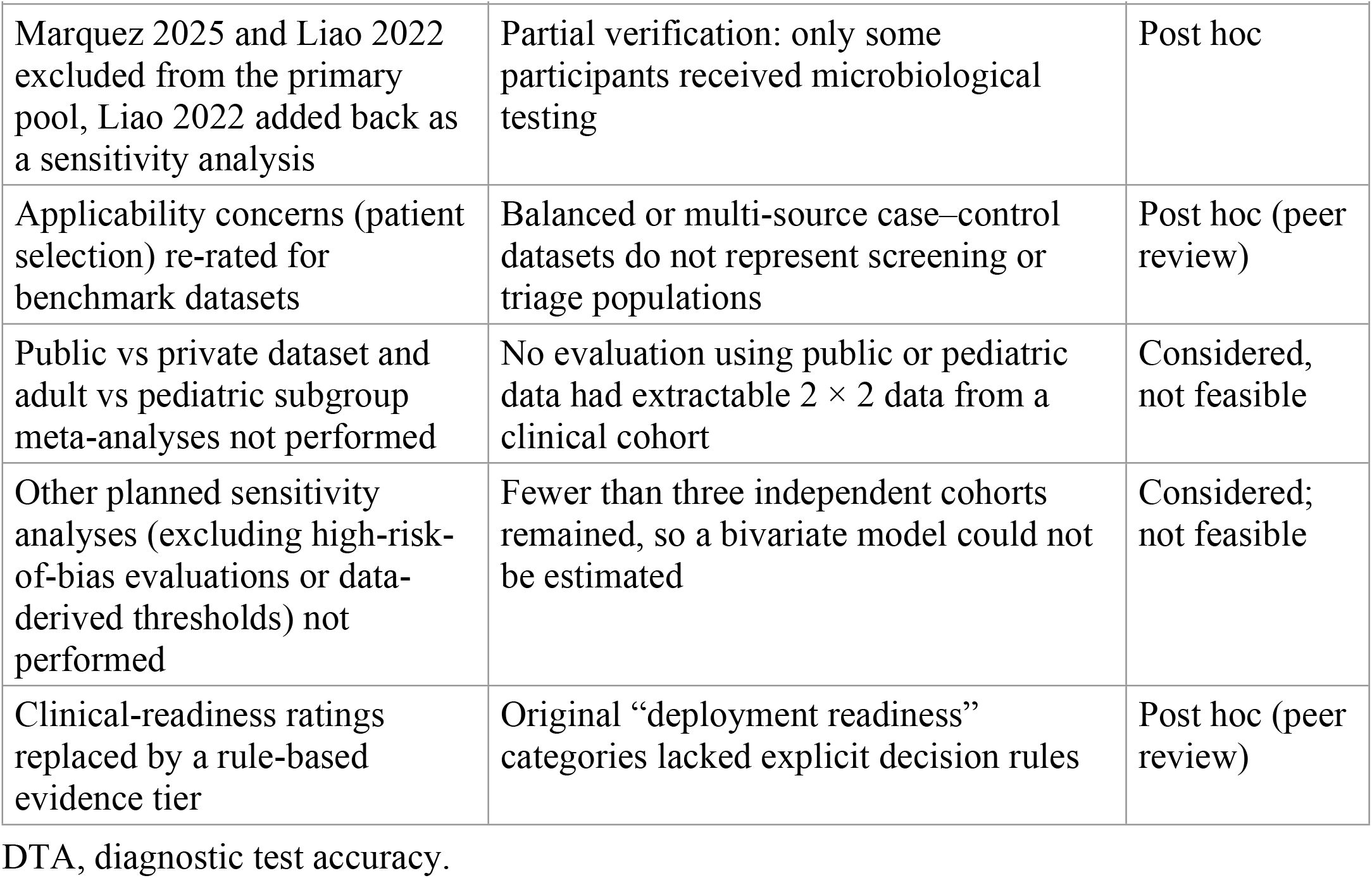
Post hoc analyses and changes made after data inspection.

### 3.2 Review framework

The review question was formulated using a PICO framework adapted for diagnostic accuracy research (Table 2). The AI algorithm was the index test and pulmonary TB was the target condition. The reference standard was recorded separately from any comparator.

**Table 2.**
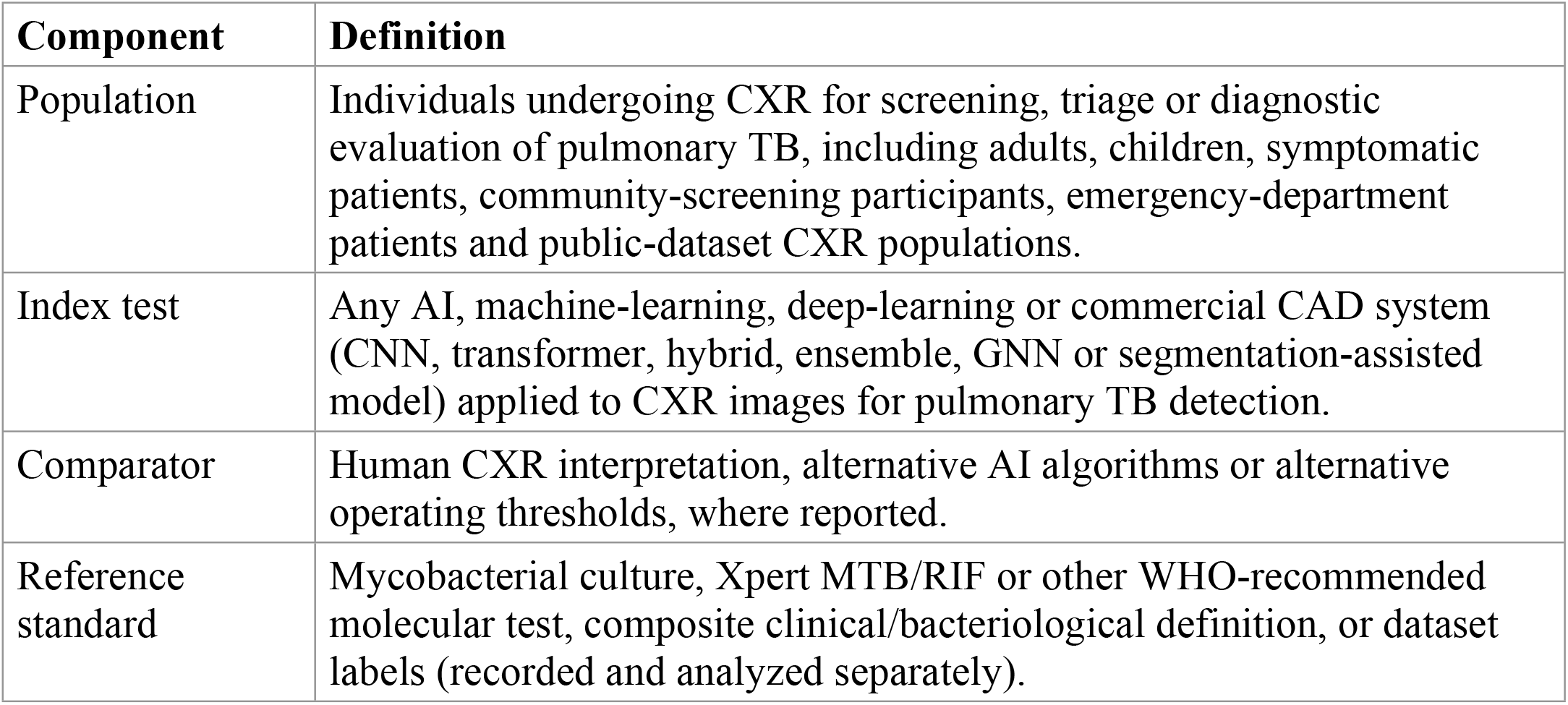
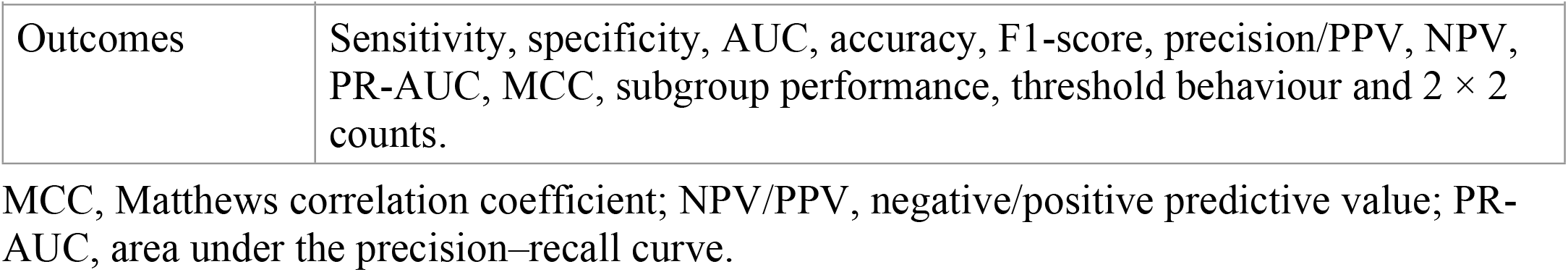
PICO framework.

| **Component** | **Definition** |
|---|---|
| Population | Individuals undergoing CXR for screening, triage or diagnostic evaluation of pulmonary TB, including adults, children, symptomatic patients, community-screening participants, emergency-department patients and public-dataset CXR populations. |
| Index test | Any AI, machine-learning, deep-learning or commercial CAD system (CNN, transformer, hybrid, ensemble, GNN or segmentation-assisted model) applied to CXR images for pulmonary TB detection. |
| Comparator | Human CXR interpretation, alternative AI algorithms or alternative operating thresholds, where reported. |
| Reference standard | Mycobacterial culture, Xpert MTB/RIF or other WHO-recommended molecular test, composite clinical/bacteriological definition, or dataset labels (recorded and analyzed separately). |
| Outcomes | Sensitivity, specificity, AUC, accuracy, F1-score, precision/PPV, NPV, PR-AUC, MCC, subgroup performance, threshold behaviour and 2 × 2 counts. |

MCC, Matthews correlation coefficient; NPV/PPV, negative/positive predictive value; PR-AUC, area under the precision–recall curve.

### 3.3 Eligibility criteria

The eligibility criteria are given in Table 3. If a study reported several algorithms or operating thresholds in the same population, the article was counted once at the study level and each algorithm (at its primary operating point) was recorded as a separate model evaluation.

**Table 3.** Inclusion and exclusion criteria.

| **Criterion** | **Definition** |
|---|---|
| Inclusion | AI, ML, DL, neural-network, commercial CAD or hybrid/ensemble model for pulmonary TB detection from CXR; reports at least one diagnostic accuracy metric or extractable 2 × 2 counts, full-text peer-reviewed journal article or conference paper in English published January 2020–March 2026. |
| Exclusion | Reviews, editorials, letters, protocols, abstracts without full text, duplicates, non-pulmonary or latent TB, non-CXR imaging, non-AI methods, animal or phantom studies, no separable pulmonary TB outcome. |

DL, deep learning; ML, machine learning.

### 3.4 Information sources and search strategy

PubMed, ScienceDirect, IEEE Xplore, OpenAlex and Lens.org were searched on 15 March 2026 for records published from January 2020 to March 2026. Only the first 100 records retrieved from Lens.org were screened. We did not have institutional access to Scopus, Web of Science and Embase, so two open scholarly indexes (OpenAlex and Lens.org), whose coverage is comparable to or broader than subscription databases, were added [39–42]. The search is therefore described as comprehensive within the available sources rather than exhaustive. After the main searches, a supplementary check in PubMed using the omitted terms (computer-aided detection/diagnosis, product names) found further clinical evaluations of commercial CAD; these were not screened for this review (Section 5.10).

The search strings were drafted with the help of a generative AI model and were run by the first author on 15 March 2026. Three concepts (tuberculosis, artificial intelligence/machine learning and chest radiography) were combined using Boolean operators, phrase searching, truncation and database-specific field tags. The exact strings executed in each database are given in Table 4 and S1 Appendix (section A). “Computer-aided detection/diagnosis” and commercial product names were not searched as separate terms; this limitation is discussed in Section 5.10.

**Table 4.** Database-specific search strings (searched 15 March 2026).

| **Database** | **Executed search string** | **Fields** | **Limits and records** |
|---|---|---|---|
| PubMed | (“Artificial Intelligence” OR “Machine Learning” OR “Deep Learning” OR AI OR ML OR DL) AND (“Tuberculosis” OR TB OR “Pulmonary Tuberculosis”) AND (detection OR diagnosis OR detect*) AND (“Chest X-ray” OR “Chest Radiograph” OR CXR) | Title, abstract, keywords (MeSH mapped) | 2020–2026; English; humans. 503 records before and 172 after filters |
| ScienceDirect | (“Artificial Intelligence” OR “Machine Learning” OR “Deep Learning”) AND (“Tuberculosis” OR TB) AND (“Chest X-ray” OR “Chest Radiograph”) | Title, abstract, keywords | 2020–2026; research articles and conference abstracts. 869 records |
| IEEE Xplore | (“Artificial Intelligence” OR “Machine Learning” OR “Deep Learning”) AND (“Tuberculosis” OR TB) AND (“Chest X-ray” OR “Chest Radiograph” OR CXR) | Metadata, abstract | No filters recorded. 573 records |
| OpenAlex | (“artificial intelligence” OR “machine learning” OR “deep learning”) AND (tuberculosis OR tb) AND (“chest x-ray” OR cxr OR “chest radiograph”) | Title, abstract | 2020–2026; English. 384 records |
| Lens.org | title/abstract:(“artificial intelligence” OR “machine learning” OR “deep learning”) AND title/abstract:(tuberculosis OR tb) AND title/abstract:(“chest x-ray” OR cxr OR “chest radiograph”) | Title, abstract | First 100 records screened |

### 3.5 Study selection

Records were merged in a spreadsheet and de-duplicated by DOI, title, author and year. The number of duplicates (44) was recorded from the consolidated file; the de-duplication procedure was not documented in detail, and the count was not independently verified. One reviewer (SAM) screened titles and abstracts, assessed full texts and extracted data using a standardized form (S1 Appendix, section B). A co-author also reviewed the work informally, but the extent of this check was not documented, so no independent second screening or extraction was recorded and inter-reviewer agreement could not be calculated. Screening and extraction were not blinded to author or journal. Reasons for exclusion at the full-text stage were recorded for only some reports (Section 4.1).

### 3.6 Data extraction

The extracted variables (Table 5) covered study characteristics, dataset details, model information, validation strategy, reference standard, diagnostic outcomes, explainability methods and limitations. Metrics that were not reported were recorded as “not reported” and not imputed. 2 × 2 counts were used only when reported or exactly derivable from reported integer counts; tables reconstructed from rounded percentages were not used. A master accounting table maps every evaluation to each analysis (S1 Appendix, section D, Table B).

**Table 5.**
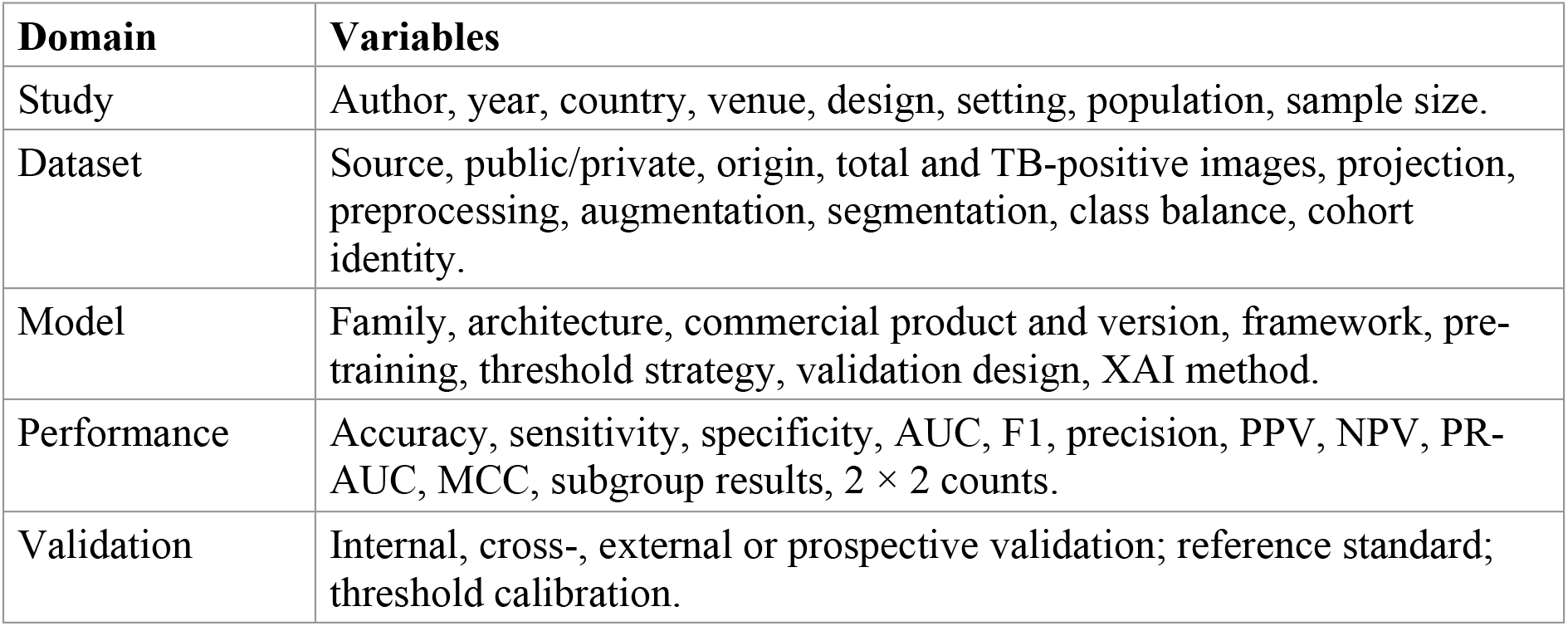
Data extraction domains.

| **Domain** | **Variables** |
|---|---|
| Study | Author, year, country, venue, design, setting, population, sample size. |
| Dataset | Source, public/private, origin, total and TB-positive images, projection, preprocessing, augmentation, segmentation, class balance, cohort identity. |
| Model | Family, architecture, commercial product and version, framework, pre-training, threshold strategy, validation design, XAI method. |
| Performance | Accuracy, sensitivity, specificity, AUC, F1, precision, PPV, NPV, PR-AUC, MCC, subgroup results, 2 × 2 counts. |
| Validation | Internal, cross-, external or prospective validation; reference standard; threshold calibration. |

### 3.7 Quality assessment

Risk of bias and applicability were assessed with QUADAS-2 [27]. Patient selection was judged on consecutive or random enrollment, avoidance of case–control sampling and artificial class balancing, and representativeness of the intended screening population. The index-test domain considered whether the AI output and threshold were prespecified and whether the threshold was derived from the test data. The reference-standard domain considered whether TB status was defined by culture, Xpert MTB/RIF or another WHO-recommended molecular test, a composite clinical definition, radiologist labels or public-dataset labels; culture or a well-defined composite definition was rated low risk, Xpert MTB/RIF alone without culture was rated unclear because a single molecular test can miss paucibacillary, culture-positive disease, and radiographic or dataset labels of uncertain provenance were rated unclear or high. Flow and timing considered whether all participants received the same reference standard, the interval between tests and exclusions after enrollment. Each domain was rated low, high or unclear. Overall risk of bias was rated high if any domain was high, low only if all four domains were low, and unclear otherwise.

Applicability concerns were rated for patient selection, index test and reference standard relative to the review question (clinical screening and triage). Risk of bias and applicability were rated by one reviewer (SAM), no independent second rating was recorded.

### 3.8 Synthesis

All included studies were synthesized narratively by model family (commercial CAD, CNN-based, transformer, hybrid/ensemble, GNN, other), setting, dataset type, population, reference standard, validation design and use of XAI. Performance was interpreted against the WHO target product profile (TPP) for a triage test (sensitivity ≥90%, specificity ≥70%) [11].

### 3.9 Statistical analysis

#### 3.9.1 Eligibility for pooling

Quantitative synthesis included evaluations from clinical cohorts with (i) complete 2 × 2 data and (ii) a microbiological or composite clinical reference standard applied to all participants. Benchmark datasets with dataset-label reference standards were excluded from the primary pool because they estimate a different target condition under case–control sampling. Marquez 2025 and Liao 2022 were excluded because of partial verification: in each, only some participants received microbiological testing (Section 4.9). A sensitivity analysis added Liao 2022 back.

#### 3.9.2 Bivariate model

Pooled sensitivity and specificity were estimated with the bivariate random-effects model of Reitsma et al. [43], which jointly models logit-transformed sensitivity and logit false-positive rate (FPR) with correlated random effects, estimated by restricted maximum likelihood (REML) using the reitsma() function of the R package mada [44]. A continuity correction of 0.5 was added to every cell (mada default, correction.control = "all").

Confidence intervals for pooled estimates are Wald intervals on the logit scale, back-transformed. The confidence interval for specificity is the back-transformed interval of the FPR. Study-level estimates in the forest plots are the observed (uncorrected) proportions with exact Clopper–Pearson 95% confidence intervals computed from the original tables. The summary ROC (SROC) curve was derived from the bivariate parameters using the Rutter-Gatsonis parameterization [45], and the area under the curve (AUC) and the normalized partial AUC restricted to the observed FPR range were computed by numerical integration.

95% prediction intervals for sensitivity and specificity were computed on the logit scale as μ^ ± *t*_k−2_ × √(SE(μ^)^2^ + τ^^2^) [38], where *k* is the number of evaluations.

#### 3.9.3 Heterogeneity

Heterogeneity within the bivariate model was quantified by the between-study standard deviations (τ) of logit sensitivity and logit FPR, their correlation (ρ), and the Zhou-Dendukuri bivariate *I*^2^ [46]; the Holling sample-size-unadjusted and -adjusted *I*^2^ ranges are reported for transparency. *I*^2^ is interpreted descriptively, as a measure of the proportion of total variability attributable to differences between evaluations on the transformed scale, not as a direct partition of clinical variation. Prediction intervals were used as the main guide for clinical interpretation.

#### 3.9.4 Non-independence of evaluations

Eight of the nine pooled evaluations shared participants with at least one other pooled evaluation: five algorithms were evaluated on the same Dhaka cohort (Qin 2021), and three were evaluated on the same prospective Karachi cohort (qXR and CAD4TB in Khan 2020; Lunit INSIGHT in Tavaziva 2022, a secondary analysis of that cohort). The nine evaluations therefore arise from three independent cohorts. The primary bivariate model treats evaluations as independent, which may understate uncertainty. We therefore conducted a post hoc analysis with one evaluation per cohort, using the first-reported evaluation in each cohort (Khan qXR, Qin qXR, Wang; *k* = 3). With three cohorts, this model is descriptive only. A multilevel model with cohort-level random effects was not fitted because three clusters cannot support it. The within-cohort sampling covariance between algorithms read on the same images could not be estimated from published data.

#### 3.9.5 Subgroup and sensitivity analyses

The planned analyses were: commercial versus custom models; public versus private datasets; internal versus external validation; adult versus pediatric; prospective versus retrospective; and exclusion of evaluations at high overall risk of bias, with dataset-label reference standards, or with data-derived thresholds. Only analyses supported by 2 × 2 data from clinical cohorts could be conducted quantitatively (Table 11); all subgroups with a single cohort or *k* ≤ 3 cohorts are descriptive and are not formal tests of effect modification. Custom models could not be pooled; commercial and custom models were therefore compared descriptively only, and no significance test was performed or is claimed.

#### 3.9.6 Diagnostic odds ratio

For comparability with earlier reviews, the diagnostic odds ratio (DOR) was pooled with the Mantel–Haenszel common-effect method and a REML random-effects model (0.5 added to tables with a zero cell). *I*^2^ for the DOR is based on Cochran’s *Q*.

### 3.10 Assessment of small-study effects

Because standard funnel plots and Egger’s regression are not valid for diagnostic accuracy data, small-study effects were to be assessed with Deeks’ funnel-plot asymmetry test [37]. The test requires at least ten evaluations (the default minimum in the R package meta); the primary pool has nine, so the test was not estimable and is not reported. Trim-and-fill was not applied.

### 3.11 Certainty of evidence

The certainty of the pooled sensitivity and specificity was rated with the GRADE approach for test accuracy [47], starting from high certainty for cohort-type accuracy studies and considering risk of bias, indirectness, inconsistency, imprecision and publication bias.

The ratings are the authors’ judgments against the WHO triage thresholds and are given with their reasons in S2 File.

### 3.12 Software and reproducibility

Analyses were run in R 4.6.0 with the packages mada and meta, and Figs 3, 4 and 7 were produced with ggplot2. The PRISMA flow diagram (Fig 1) and Figs 2, 5 and 6 were drawn in Python (matplotlib) from the same extracted data and model outputs. The R scripts and extraction data are provided in S1 File. The first author ran all analyses, checked every output and takes responsibility for the content.

**Fig 1.**
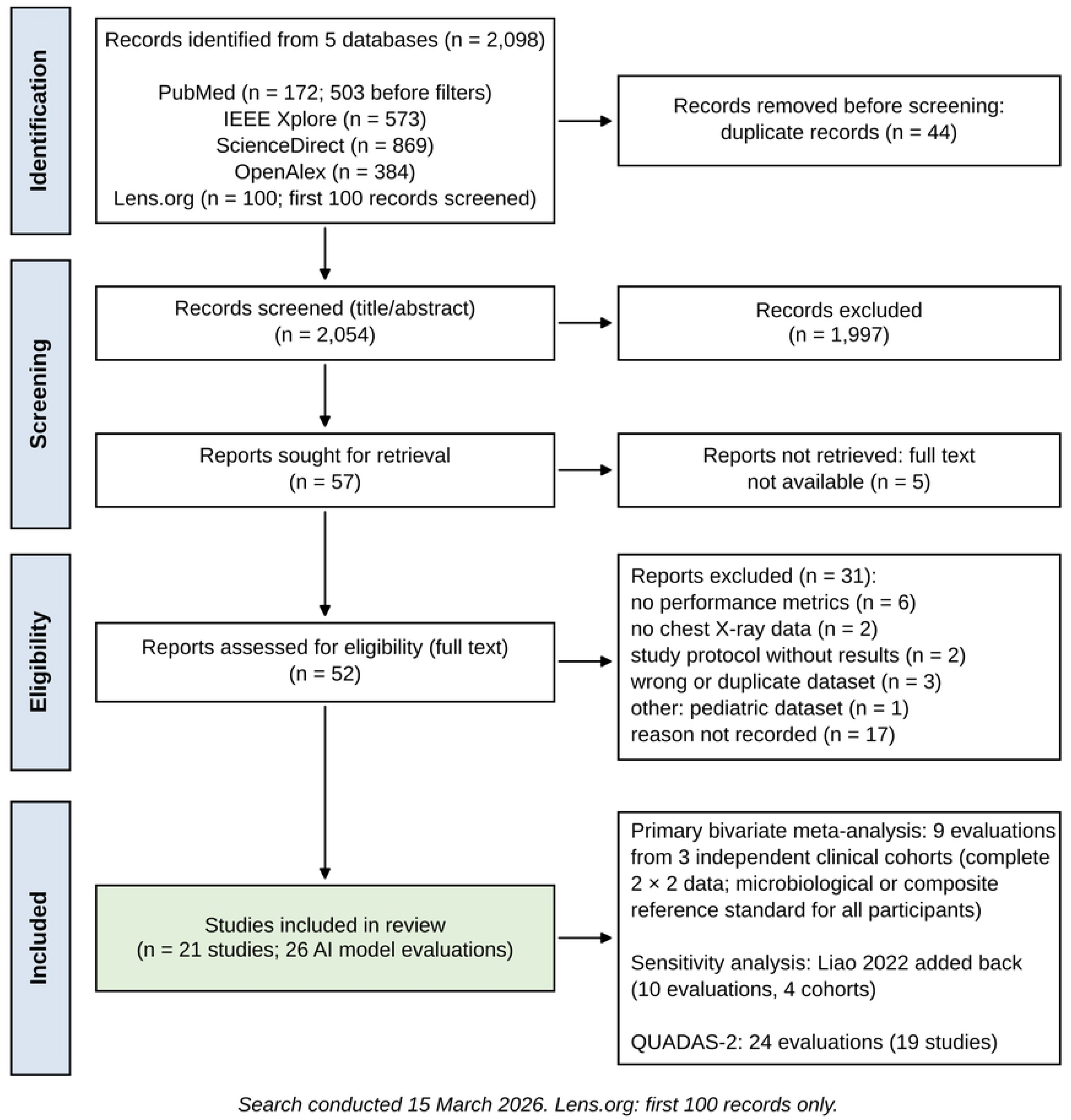
PRISMA 2020 flow diagram of study identification, screening and inclusion. The nine pooled evaluations originate from three independent clinical cohorts (Karachi, Pakistan; Dhaka, Bangladesh; Taipei, Taiwan); Liao 2022 (Yingde, China) was analyzed in a sensitivity analysis only. QUADAS-2 was applied to 24 evaluations from 19 studies. AI, artificial intelligence.

**Fig 2.**
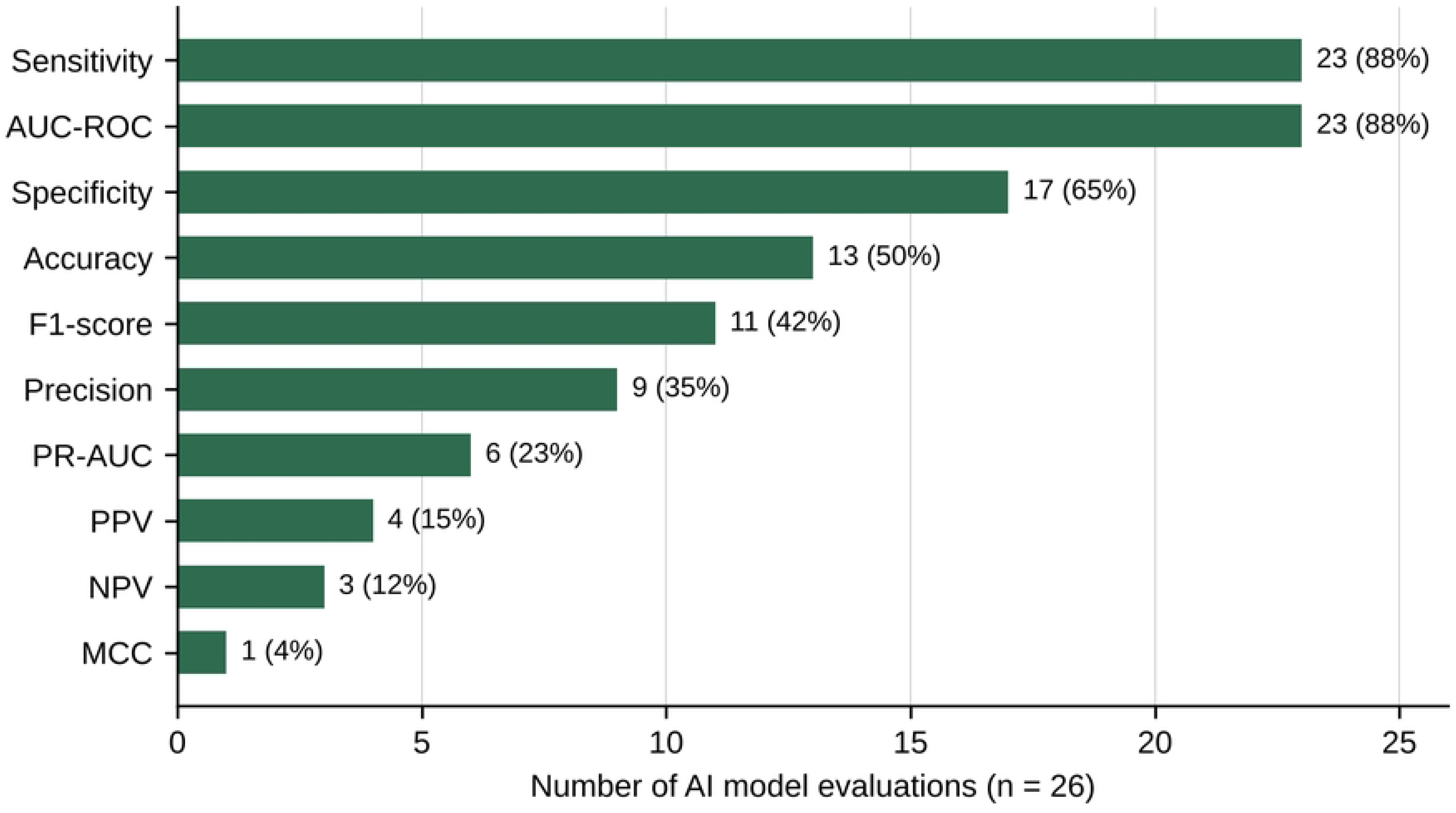
Number of AI model evaluations reporting each diagnostic performance metric (n = 26). Percentages are of the 26 evaluations. AUC-ROC, area under the receiver operating characteristic curve; MCC, Matthews correlation coefficient; NPV/PPV, negative/positive predictive value; PR-AUC, area under the precision–recall curve.

**Fig 3.**
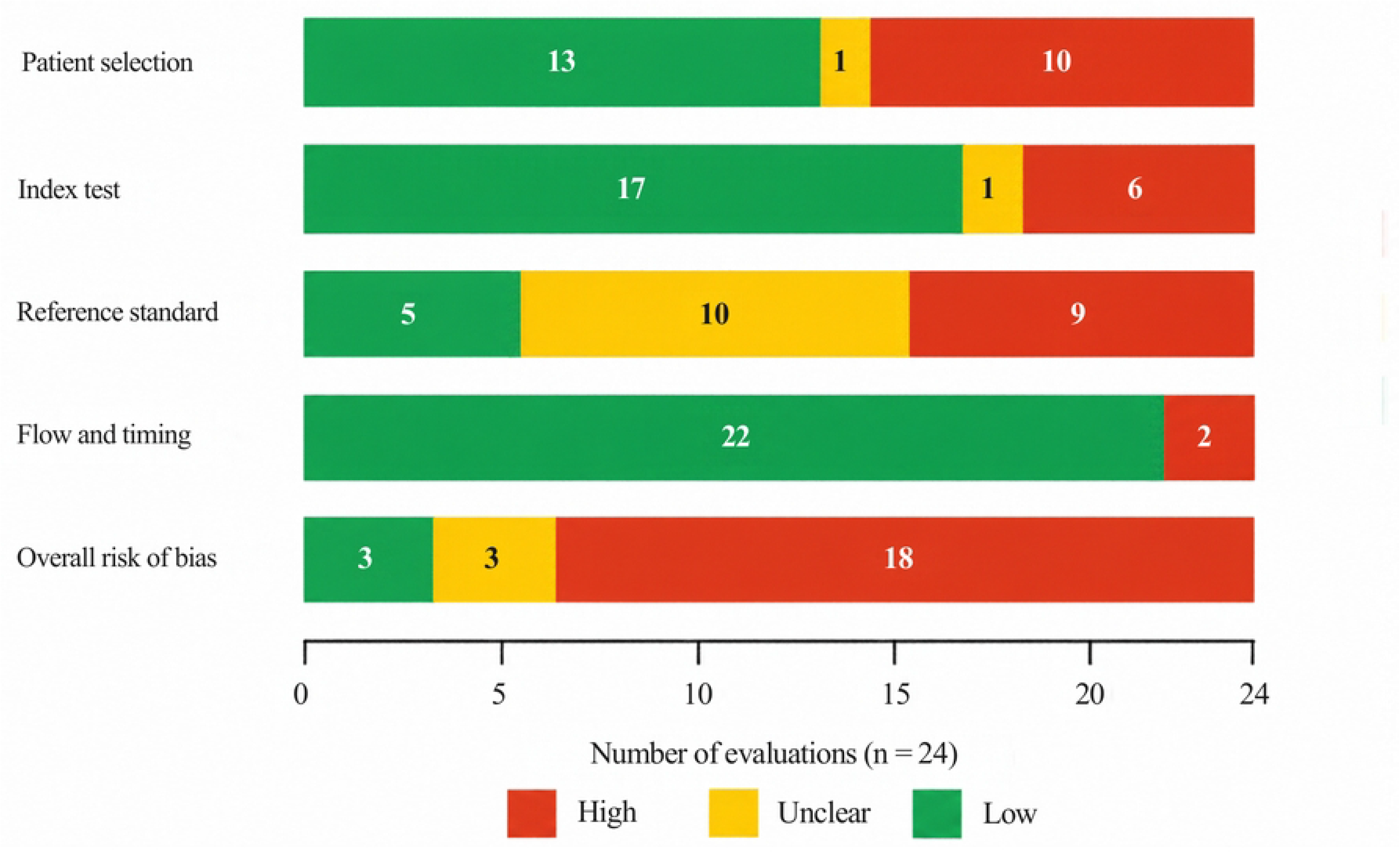
QUADAS-2 summary for the 24 assessed evaluations. Risk of bias by domain and overall (high if any domain high; low only if all domains low; otherwise unclear). Per-evaluation judgements are shown in Table 7 and in Fig A in S1 Appendix.

**Fig 4.**
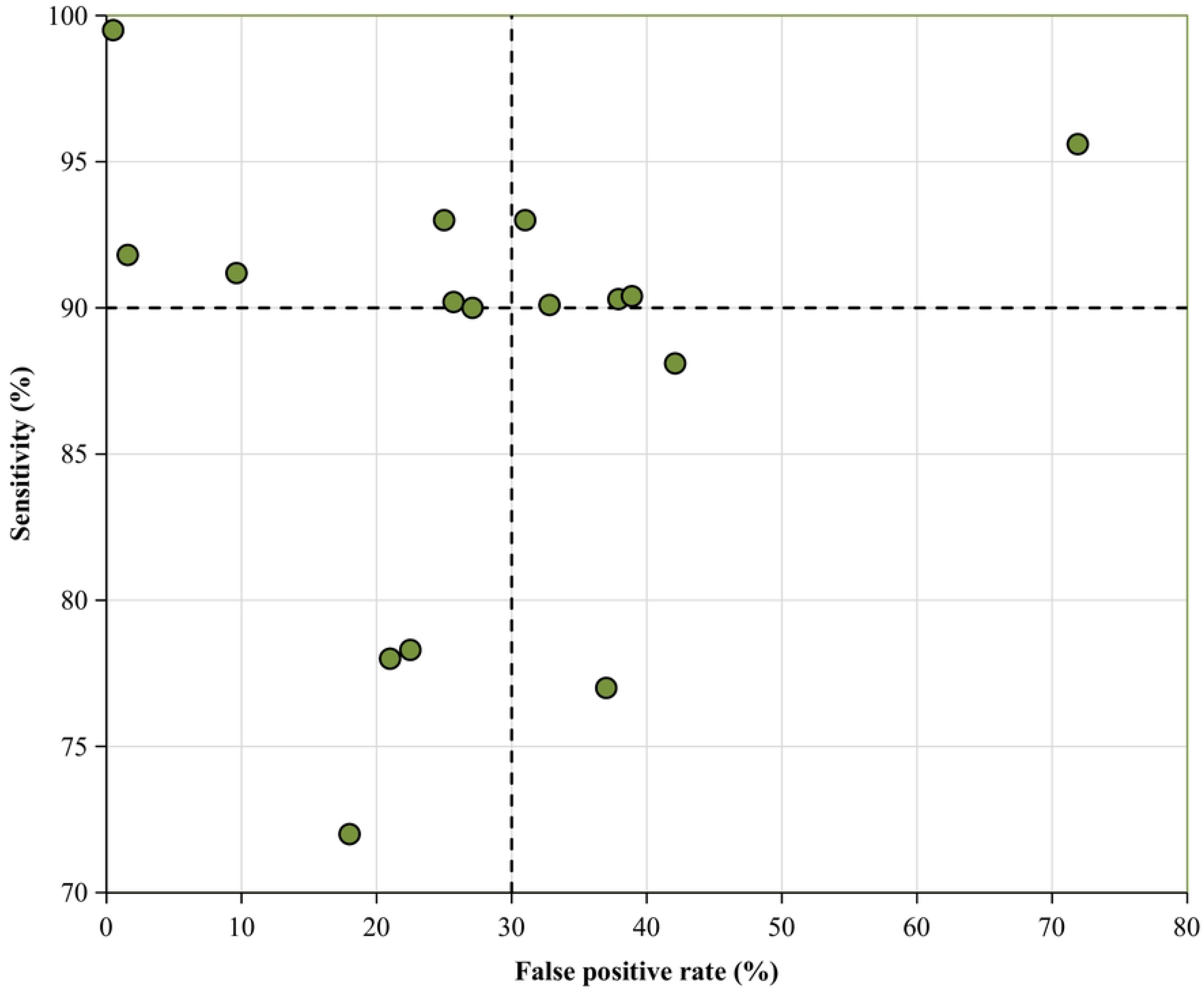
ROC-space distribution of 16 AI model evaluations reporting both sensitivity and specificity. Seventeen evaluations reported both measures; Liao 2022 is omitted because of partial verification, and nine evaluations did not report specificity. Each point is one evaluation; the values are given in Table 8. Dashed lines mark the WHO TPP for a triage test (sensitivity ≥90%, specificity ≥70%, i.e. false-positive rate ≤30%). Qin 2021 algorithms are plotted at the 90%-sensitivity operating point; Tavaziva 2022 at score threshold 15.

**Fig 5.**
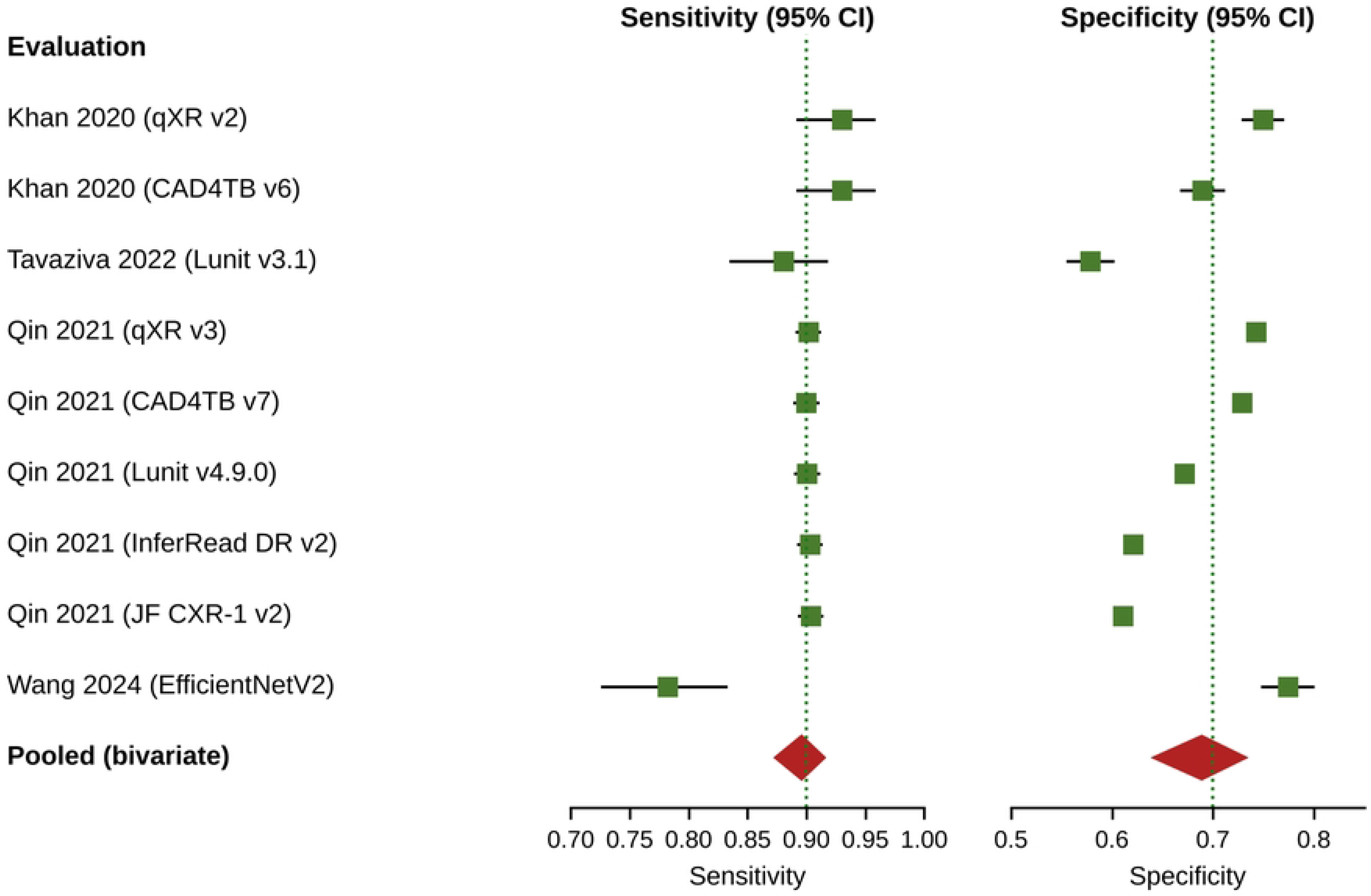
Forest plot of sensitivity (left) and specificity (right) for the nine pooled evaluations. One row per evaluation, in the order of Table 10; Khan 2020 and Tavaziva 2022 share the Karachi cohort. Squares show observed estimates; whiskers are exact Clopper– Pearson 95% CIs (some are narrower than the markers). Red diamonds show the bivariate random-effects pooled estimate with its 95% CI. Dotted green lines mark the WHO TPP thresholds (sensitivity 90%; specificity 70%). Zhou–Dendukuri *I*^2^ ≈ 75%; τ (logit sensitivity) = 0.31, τ (logit FPR) = 0.32, ρ = 0.30.

**Fig 6.**
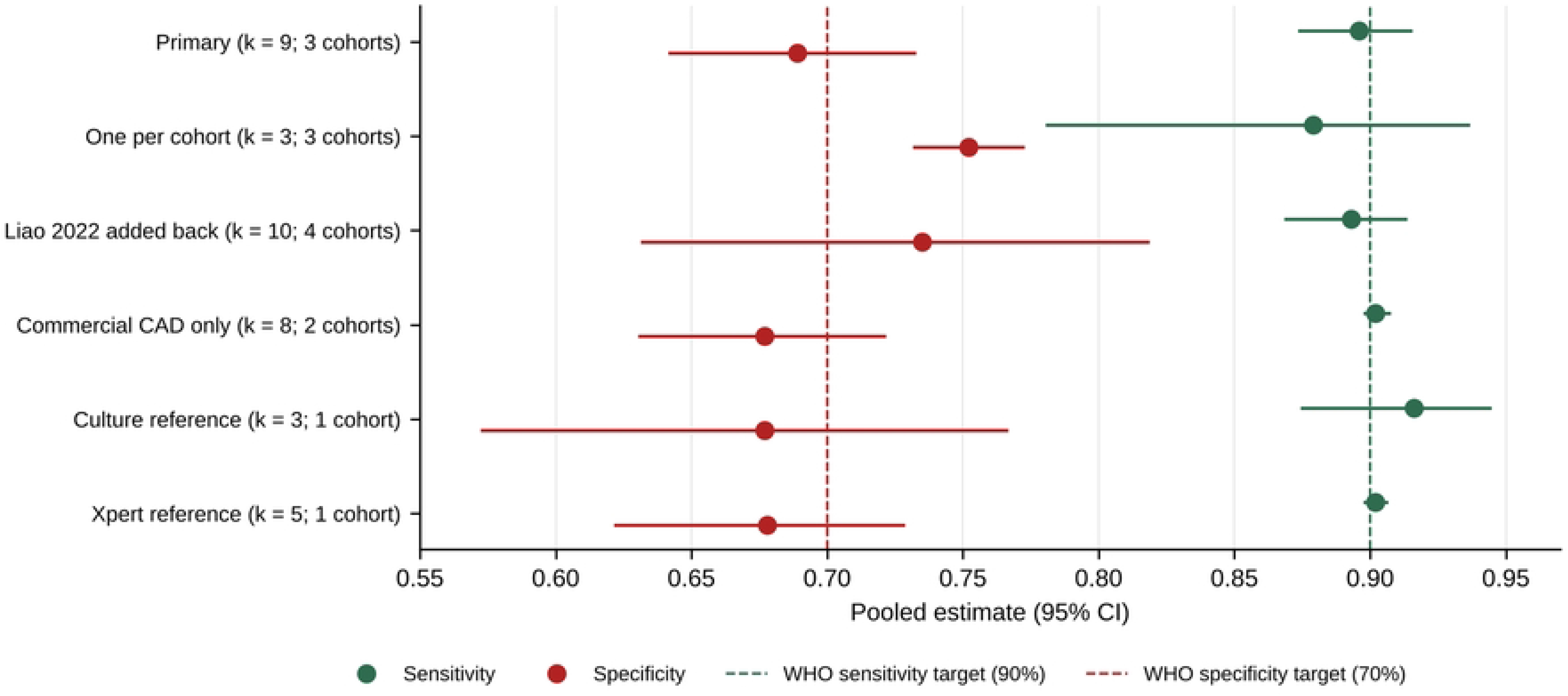
Pooled sensitivity and specificity across the primary, dependence-adjusted, sensitivity and subgroup analyses. Points are pooled estimates and horizontal lines their 95% CIs (values in Table 11). Dashed vertical lines mark the WHO TPP thresholds (green, sensitivity 90%; red, specificity 70%). Analyses with a single cohort or *k* ≤ 3 cohorts are descriptive.

**Fig 7.**
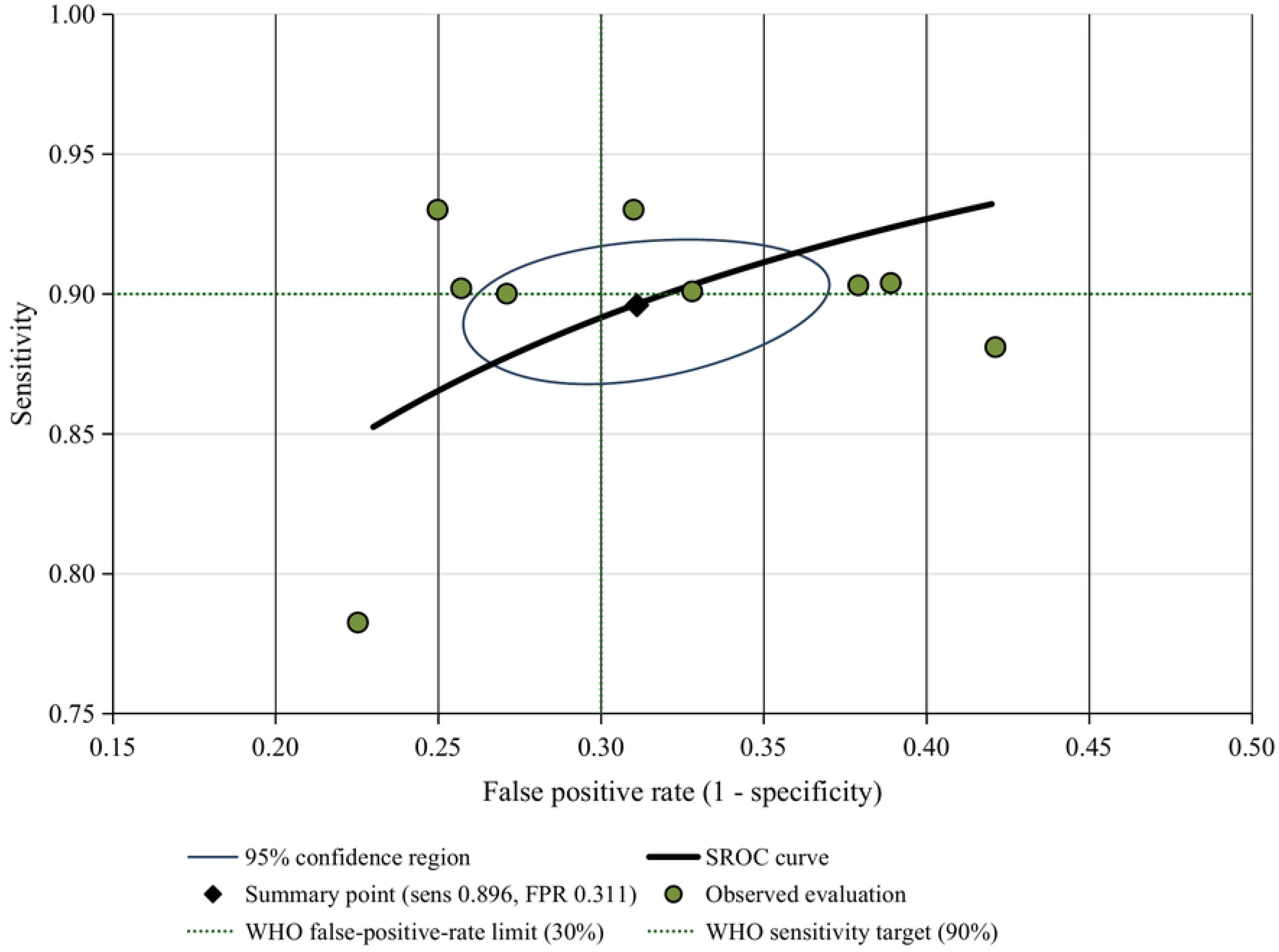
Summary ROC curve from the bivariate model for the nine pooled evaluations. Rutter–Gatsonis parameterization, drawn over the observed FPR range, with the summary point (sensitivity 0.896, specificity 0.689) and its 95% confidence region. Filled circles show the individual evaluations; dotted lines mark the WHO triage thresholds (sensitivity 90%, false-positive rate 30%). AUC 0.882; normalized partial AUC 0.889.

**Table 6.** Datasets used by the included studies (as reported by study authors).

| **Dataset** | **Origin** | **Access** | **Images** | **TB-positive** | **Studies using it** |
|---|---|---|---|---|---|
| Shenzhen | China | Public | 662 | 336 | 6 [10, 15–17, 20, 25] |
| Montgomery County | USA | Public | 138 | 58 | 5 [10, 15, 16, 20, 25] |
| TBX11K | China | Public | 11,200 | 800 | 1 [10] |
| NIH ChestX-ray14/8 | USA | Public | 112,120 | None (no TB label; non-TB source) | 2 [17, 25] |
| Belarus TB portal | Belarus | Public | 304 | 304 | 2 [10, 20] |
| NIAID TB portal | Multi | Public | 3,000 | 3,000 | 2 [10, 16] |
| RSNA pneumonia | USA | Public | Subset | None (non-TB source) | 2 [10, 20] |
| TB Chest X-ray Database (Kaggle) | Multi | Public | 4,200 | 700 | 5 [12, 15, 18, 21, 23] |
| Other Kaggle compilations<sup>a</sup> | Multi | Public | 600–12,841 | 300–3,495 | 3 [14, 22, 24] |
| IN-CXR (ICMR-NIRT survey) | India | Public | 22,406<sup>b</sup> | 11,203 abnormal<sup>b</sup> | 1 [13] |
| DA/DB | India | Private | 153 | 153 | 1 [10] |
| Indus Hospital cohort | Pakistan | Private | 2,198 | 272 (culture) | 2 [3, 7] |
| Dhaka screening centres | Bangladesh | Private | 23,954 | 3,675 (Xpert) | 1 [4] |
| TBIHSS case finding | Philippines | Private | 5,740 | 749 (mWRD) | 1 [6] |
| NTUH | Taiwan | Private | 3,498 | 253 | 1 [25] |
| Yingde mobile screening | China | Private | 2,543 | 8 | 1 [5] |
| CISM / pTBred / CNH-TB-PEDs | Mozambique / Spain / USA | Private | 218 / 565 / 71 | 38 / 253 / 19 | 1 [8] |

<sup>a</sup> CXR Masks and Labels (600 images; 300 TB), TBC dataset (790; 395) and Pneumonia–TB compilation (12,841; 3,495). <sup>b</sup> After oversampling to a 1:1 ratio; “abnormal” is survey-read radiographic abnormality, not confirmed TB. mWRD, WHO-recommended molecular rapid diagnostic.

**Table 7.** QUADAS-2 risk of bias and applicability judgements (L, low; H, high; U, unclear).

| **Evaluation** | **RoB PS** | **RoB IT** | **RoB RS** | **RoB FT** | **RoB overall** | **Appl. PS** | **Appl. IT** | **Appl. RS** |
|---|---|---|---|---|---|---|---|---|
| Acharya 2022 | L | L | U | L | U | H | L | L |
| Bhandari 2022 | L | L | U | L | U | H | L | L |
| Khan 2020 (qXR) | L | L | L | L | L | L | L | L |
| Khan 2020 (CAD4TB) | L | H | L | L | H | L | L | L |
| Wang 2024 | H | L | L | L | H | L | L | L |
| Liao 2022 | U | L | H | H | H | L | L | L |
| Qin 2021 (qXR) | L | H | U | L | H | L | L | L |
| Qin 2021 (CAD4TB) | L | H | U | L | H | L | L | L |
| Qin 2021 (Lunit) | L | H | U | L | H | L | L | L |
| Qin 2021 (InferRead) | L | H | U | L | H | L | L | L |
| Qin 2021 (JF CXR-1) | L | H | U | L | H | L | L | L |
| Capellán-Martín 2025 | L | L | L | L | L | L | L | L |
| Siddharth 2025 | L | L | U | L | U | H | L | L |
| Maashi 2025 | H | L | U | L | H | H | L | L |
| Sharmila 2025 | H | U | H | L | H | H | L | L |
| Tavaziva 2022 | L | L | L | L | L | L | L | L |
| Sharma 2024 | H | L | H | L | H | H | L | L |
| Sathitratanacheewin 2020 | H | L | H | L | H | H | L | L |
| Mirugwe 2025 | H | L | H | L | H | H | L | L |
| Marquez 2025 | L | L | H | H | H | L | L | L |
| Pala 2025 | H | L | H | L | H | H | L | L |
| Shahzad 2025 | H | L | H | L | H | H | L | L |
| Ridwan 2026 | H | L | H | L | H | H | L | L |
| Akbari 2023 | H | L | U | L | H | H | L | L |
| **Totals (L/U/H)** | 13/1/10 | 17/1/6 | 5/10/9 | 22/0/2 | 3/3/18 | 12/0/12 | 24/0/0 | 24/0/0 |

RoB, risk of bias; Appl., applicability concern; PS, patient selection; IT, index test; RS, reference standard; FT, flow and timing.

**Table 8.**
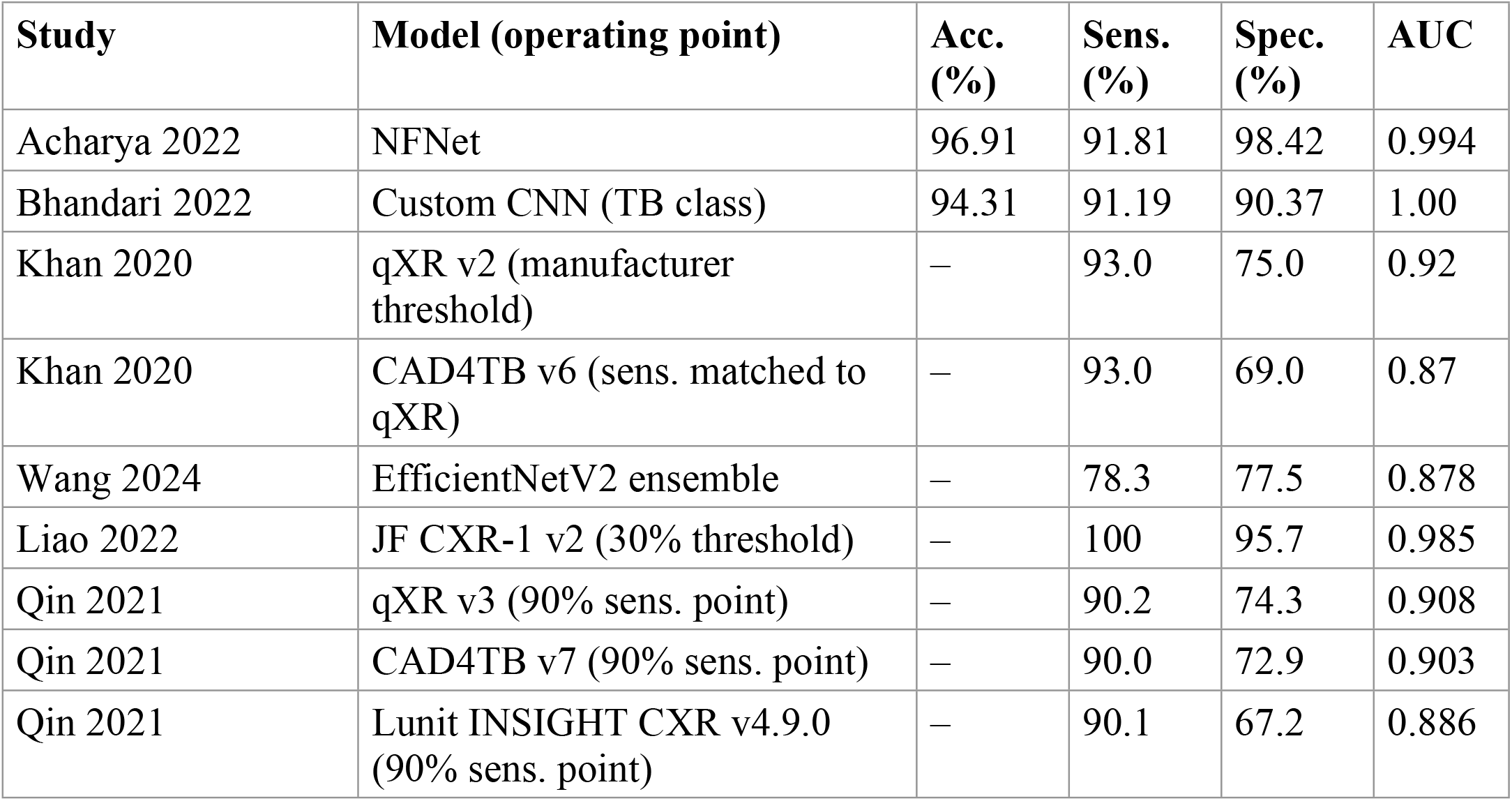
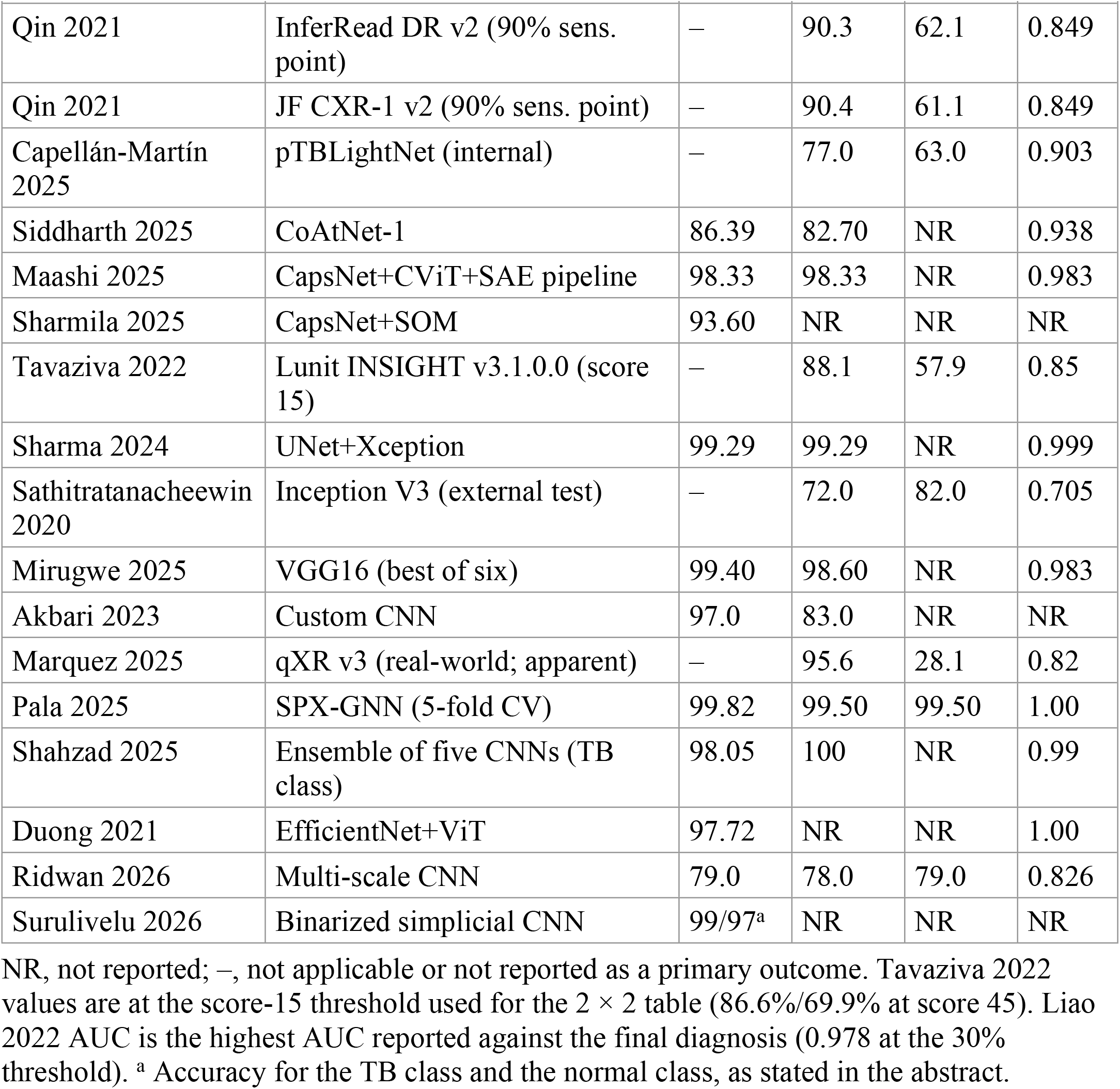
Reported diagnostic accuracy of the 26 AI model evaluations.

**Table 9.** Explainable AI methods and clinical validation.

| **Study** | **XAI method** | **Clinical validation** |
|---|---|---|
| Bhandari 2022 [12] | SHAP, LIME, Grad-CAM | Two radiologists; heatmaps aligned with TB findings |
| Wang 2024 [25] | Grad-CAM | Visualization only |
| Siddharth 2025 [13] | LIME (superpixel) | Visualization only |
| Pala 2025 [23] | Perturbation-based superpixel importance | Visualization only |
| Shahzad 2025 [24] | Grad-CAM | Visualization only |

**Table 10.** 2 × 2 data used in the primary quantitative synthesis, with cohort and reference standard.

| **Evaluation** | **Cohort** | **Reference** | **RoB** | **TP** | **FN** | **FP** | **TN** | **Sens.** | **Spec.** |
|---|---|---|---|---|---|---|---|---|---|
| Khan 2020 (qXR v2) | Karachi | Culture | Low | 253 | 19 | 481 | 1,445 | 0.930 | 0.750 |
| Khan 2020 (CAD4TB v6) | Karachi | Culture | High | 253 | 19 | 597 | 1,329 | 0.930 | 0.690 |
| Tavaziva 2022 (Lunit v3.1) | Karachi | Culture | Low | 237 | 32 | 809 | 1,112 | 0.881 | 0.579 |
| Qin 2021 (qXR v3) | Dhaka | Xpert | High | 3,315 | 360 | 5,212 | 15,067 | 0.902 | 0.743 |
| Qin 2021 (CAD4TB v7) | Dhaka | Xpert | High | 3,308 | 367 | 5,496 | 14,783 | 0.900 | 0.729 |
| Qin 2021 (Lunit v4.9.0) | Dhaka | Xpert | High | 3,311 | 364 | 6,651 | 13,628 | 0.901 | 0.672 |
| Qin 2021 (InferRead DR v2) | Dhaka | Xpert | High | 3,319 | 356 | 7,686 | 12,593 | 0.903 | 0.621 |
| Qin 2021 (JF CXR-1 v2) | Dhaka | Xpert | High | 3,322 | 353 | 7,888 | 12,391 | 0.904 | 0.611 |
| Wang 2024 (EfficientNetV2) | NTUH | Composite | High | 198 | 55 | 248 | 853 | 0.783 | 0.775 |

RoB, overall QUADAS-2 risk of bias. Khan 2020 and Tavaziva 2022 analyzed the same prospective Karachi cohort. Liao 2022 (Yingde; composite reference; partial verification; TP 8, FN 0, FP 110, TN 2,425) was analyzed only in the sensitivity analysis.

**Table 11.** Composition and results of the dependence-adjusted, sensitivity and subgroup analyses (bivariate model).

| **Analysis** | **Evaluations included** | **k (cohorts)** | **Sensitivity (95% CI)** | **Specificity (95% CI)** |
|---|---|---|---|---|
| Primary | All nine in Table 10 | 9 (3) | 0.896 (0.874–0.915) | 0.689 (0.642–0.732) |
| One per cohort (post hoc) | Khan qXR, Qin qXR, Wang | 3 (3) | 0.879 (0.781–0.936) | 0.752 (0.732–0.772) |
| Liao 2022 added back (post hoc) | All nine + Liao 2022 | 10 (4) | 0.893 (0.869–0.913) | 0.735 (0.632–0.818) |
| Commercial CAD only | All except Wang | 8 (2) | 0.902 (0.898–0.907) | 0.677 (0.631–0.721) |
| Culture reference | Khan qXR, Khan CAD4TB, Tavaziva | 3 (1) | 0.916 (0.875–0.944) | 0.677 (0.573–0.766) |
| Xpert reference | Five Qin 2021 algorithms | 5 (1) | 0.902 (0.898–0.906) | 0.678 (0.622–0.728) |

None of the pooled evaluations used dataset-label reference standards, so the planned sensitivity analysis excluding dataset-label studies is identical to the primary analysis. Estimates with *k* ≤ 3 cohorts or a single cohort are imprecise and descriptive. The Xpert-reference analysis uses the same cohort as most of the commercial-CAD analysis.

## 4 Results

### 4.1 Study selection

The searches retrieved 2,098 records (PubMed 172 after filters, from 503 before filters; IEEE Xplore 573, ScienceDirect 869, OpenAlex 384, Lens.org 100, first 100 records only). After removal of 44 duplicates, 2,054 records were screened on title and abstract and 1,997 were excluded. Fifty-seven reports were sought for retrieval; the full text of five was not available, and 52 reports were assessed in full text. Thirty-one were excluded (Fig 1), and 21 studies with 26 AI model evaluations were included. Reasons for exclusion at the full-text stage were recorded for 14 of the 31 reports: no performance metrics (6), no chest X-ray data (2), study protocol without results (2), wrong or duplicate dataset (3) and a pediatric dataset (1); no reason was recorded for the other 17.

### 4.2 Characteristics of included studies

#### 4.2.1 Geography

The 21 studies were published in 2020–2026 and involved 21 countries in Asia, Africa, Europe, North America and Oceania (country of data origin or author affiliation; one study did not state a country [19]). India contributed four studies and Pakistan three, and five studies were multi-country collaborations [8, 12, 18, 20, 24]. Most of the output came from high-burden countries in South and Southeast Asia.

#### 4.2.2 Study design

Three studies recruited participants prospectively [3, 5, 7]. Khan 2020 and Tavaziva 2022 studied the same prospective Karachi cohort, in which every participant received culture, whereas Liao 2022 applied the reference standard to only a subset of participants (partial verification). Marquez 2025 was a retrospective analysis of case-finding program data in which only CAD-positive or symptom-positive participants received confirmatory testing (also partial verification) [6]. Three were retrospective clinical cohort studies [4, 8, 25]. Five were cross-sectional analyses of pre-existing public or survey datasets [10, 12–14, 17], and nine were algorithm-development studies without clinical validation, one of them a conference paper [15, 16, 18–24].

#### 4.2.3 Populations

Three studies enrolled symptomatic adults in triage or diagnostic settings [3, 4, 7]; two involved community screening or active case finding [5, 6]; and one used a national TB prevalence survey with radiographic readings [13]. One was set in an emergency department [25], and one was exclusively pediatric [8]. The remaining 13 used public or institutional image collections without clinical characterization. HIV status was reported for only a minority of clinical cohorts (99.5% HIV-negative in the Karachi cohort [3, 7]), and no study reported performance stratified by HIV status.

#### 4.2.4 Datasets

Six studies used only private clinical cohorts [3–8]; 14 used public datasets alone or combined with private data; and one did not specify its data source [19]. Table 6 lists the datasets. Sample sizes ranged from 71 images (CNH-TB-PEDs) to 112,120 (NIH ChestX-ray14), and clinical cohorts from 71 to 23,954 participants. Several public compilations (e.g. the Kaggle TB Chest X-ray Database) themselves aggregate Shenzhen, Montgomery, Belarus and NIAID images, so overlap between “different” public datasets is likely.

#### 4.2.5 Model families

The 26 evaluations comprised commercial CAD systems (10: qXR v2/v3, CAD4TB v6/v7, Lunit INSIGHT CXR v3.1/v4.9, InferRead DR v2, JF CXR-1 v2 in Liao 2022 and Qin 2021, and qXR v3 in Marquez 2025); CNN-based models (8: NFNet, two custom CNNs, EfficientNetV2, UNet+Xception, Inception V3, VGG16, multi-scale CNN); transformer-based models, including convolution–transformer hybrids (2: CoAtNet-1, EfficientNet+ViT); other hybrid or ensemble architectures (4: pTBLightNet, a CapsNet+CViT+SAE pipeline, CapsNet+SOM and a five-CNN ensemble); one GNN (SPX-GNN); and one other model (binarized simplicial CNN), totaling 26.

#### 4.2.6 Reference standards

Reference standards were mycobacterial culture (2 studies) [3, 7], Xpert MTB/RIF or another WHO-recommended molecular test (2) [4, 6], a composite bacteriological/clinical definition (3) [5, 8, 25], dataset labels assigned by dataset curators or survey teams (13) [10, 12–18, 20–24], and not reported with sufficient clarity to categorize (1) [19], total 21. Only the seven studies in the first three categories define the target condition in a way that matches clinical practice.

### 4.3 Reporting of performance metrics

Sensitivity was extractable for 23 of 26 evaluations (88%) and specificity for 17 (65%); these 17 are the evaluations reporting both measures (Fig 4). Three evaluations reported neither: Sharmila [15] (accuracy and F1-score only), Duong [20] (accuracy and AUC only) and Surulivelu [19] (accuracy only: 99% for the TB class and 97% for the normal class). AUC was reported for 23 evaluations (88%), accuracy for 13 (50%), F1-score for 11 (42%), precision for 9 (35%), PR-AUC for 6 (23%), PPV for 4 (15%), NPV for 3 (12%) and MCC for 1 (4%) (Fig 2). A complete 2 × 2 table was reported by only 11 evaluations (42%). No study reported calibration.

### 4.4 Risk of bias and applicability

QUADAS-2 was applied to 24 of the 26 evaluations (19 of 21 studies); Duong [20] and Surulivelu [19] described their data and reference standards too briefly to support domain-level judgments. Domain tallies (Table 7; Fig 3; Fig A in S1 Appendix) were: patient selection 13 low, 10 high, 1 unclear; index test 17 low, 6 high, 1 unclear; reference standard 5 low, 9 high, 10 unclear; flow and timing 22 low and 2 high (Marquez 2025 and Liao 2022, partial verification).

According to the decision rule (high if any domain high; low only if all domains low), overall risk of bias was high in 18 evaluations (75.0%), unclear in 3 (12.5%) and low in only 3 (12.5%): Khan 2020 (qXR), Tavaziva 2022 and Capellán-Martín 2025. High patient-selection risk arose mainly from artificially balanced or multi-source benchmark datasets and from retrospective case–control sampling; high index-test risk from thresholds chosen on the evaluation data (all five Qin 2021 algorithms, which were compared at a fixed 90%-sensitivity operating point, and CAD4TB in Khan 2020); high reference-standard and flow-and-timing risk from partial verification (Liao 2022, Marquez 2025) and from dataset or survey labels; and unclear reference-standard risk from reliance on Xpert MTB/RIF without culture or on labels of uncertain provenance.

*Applicability.* Concerns regarding patient selection were high for 12 evaluations whose data are not representative of a screening or triage population: five used approximately balanced (about 50% TB) datasets (Sharma 2024, Sathitratanacheewin 2020, Ridwan 2026, Siddharth 2025, Maashi 2025), and seven used multi-source compilations in which TB and non-TB images derive from different populations (Acharya 2022, Bhandari 2022, Sharmila 2025, Mirugwe 2025, Akbari 2023, Pala 2025, Shahzad 2025). The other 12 had low concern. Index-test and reference-standard applicability concerns were judged low for all 24.

### 4.5 Diagnostic accuracy of AI models

Table 8 summarizes the reported performance. Sensitivity ranged from 72% (Inception V3 on external data) to 100% (JF CXR-1 at a 30% threshold; five-CNN ensemble for the TB class), specificity from 28.1% (qXR v3 in Philippine case finding) to 99.5% (SPX-GNN), and AUC from 0.705 to 1.00.

### 4.6 Performance by model family

#### 4.6.1 Commercial CAD systems

Five studies reported ten evaluations of commercial CAD. In the prospective Karachi cohort, qXR v2 had 93% sensitivity and 75% specificity (AUC 0.92) at the manufacturer’s prespecified threshold, and CAD4TB v6 93% and 69% (AUC 0.87) at a sensitivity-matched threshold [3]; Lunit INSIGHT v3.1 on the same cohort had 88.1% and 57.9% (AUC 0.85) [7]. In Dhaka, at a common 90%-sensitivity operating point, specificity was 74.3% for qXR v3, 72.9% for CAD4TB v7, 67.2% for Lunit v4.9.0, 62.1% for InferRead DR v2 and 61.1% for JF CXR-1 v2 [4]. In Chinese community screening, JF CXR-1 identified all eight TB cases at a 30% threshold with 95.7% specificity (sensitivity 95% CI 63–100%), but not all participants underwent microbiological testing [5]. In Philippine case finding, qXR v3 had 95.6% apparent sensitivity and 28.1% apparent specificity at a fixed threshold [6], which shows the gap between controlled and operational settings.

#### 4.6.2 CNN-based models

NFNet achieved 96.9% accuracy and an AUC of 0.994 [10]. The deep-learning model of Bhandari et al. reached 94.3% accuracy, with 91.2% sensitivity and 90.4% specificity for the TB class [12]. EfficientNetV2 had 78.3% sensitivity and 77.5% specificity (AUC 0.878; 0.94 for PA views) in an emergency-department cohort [25]. The multi-scale CNN reached 79% accuracy and 78% sensitivity (AUC 0.826) [22]; a custom CNN 97% accuracy but 83% sensitivity [21]; and VGG16 99.4% accuracy with 98.6% sensitivity [18]. Inception V3 dropped to 72% sensitivity and 82% specificity (AUC 0.705) on an external dataset [17].

#### 4.6.3 Transformer-based and hybrid models

CoAtNet-1 reached 86.4% accuracy and an AUC of 0.938 on IN-CXR [13], and an EfficientNet–ViT hybrid 97.7% accuracy and an AUC of 1.00 [20]. A multi-stage ResUNet/EfficientNetV2/CapsNet/CViT/SAE pipeline achieved 98.3% accuracy on a balanced dataset of 600 images [14]; CapsNet+SOM reached 93.6% accuracy (framed as multidrug-resistant TB detection but trained on public TB-versus-normal datasets) [15]; and a five-CNN ensemble reached 98.1% accuracy with 100% TB recall [24]. pTBLightNet had an internal AUC of 0.903, which fell to 0.682 on external low-burden pediatric data [8]. The data do not allow conclusions about the relative resource requirements of transformer models.

#### 4.6.4 Graph neural networks and other models

SPX-GNN reached 99.8% accuracy and an AUC of 1.00 under five-fold cross-validation on the Kaggle TB Chest X-ray Database [23]. Because it was evaluated on a different, curated dataset, this result cannot be compared directly with estimates from clinical cohorts and needs external validation. A binarized simplicial CNN reported 99% accuracy for the TB class and 97% for the normal class, without sensitivity or specificity [19].

### 4.7 Explainable AI

Five studies used XAI (Table 9). Only Bhandari et al. validated the explanations formally, with two radiologists confirming that highlighted regions corresponded to TB-compatible abnormalities [12]; the others presented visualizations without clinical validation.

### 4.8 Distribution of performance in ROC space

Seventeen evaluations reported both sensitivity and specificity; the other nine did not report specificity. Fig 4 plots 16 of the 17; Liao 2022 (100% sensitivity and 95.7% specificity at a 30% threshold) is omitted because of partial verification. Six of the 16 plotted evaluations met both WHO TPP thresholds at the reported operating point: three clinical-cohort evaluations of commercial CAD (Khan qXR; Qin qXR and CAD4TB) and three models evaluated internally on benchmark datasets (Acharya 2022, Bhandari 2022, Pala 2025); Liao 2022 also met both thresholds. Among the other commercial evaluations the false-positive rate was 31–42% in triage cohorts and 72% in real-world case finding [6].

Models evaluated on external or clinical data performed more modestly: 78.3%/77.5% for EfficientNetV2 [25], 72%/82% for Inception V3 on external data [17] and 77%/63% for the pediatric model [8]. No single algorithm version has been shown to meet both thresholds in more than one setting with complete verification, qXR met them in Karachi (v2, at the manufacturer threshold) and in Dhaka (v3, at an operating point derived from the test data).

### 4.9 Meta-analysis of diagnostic accuracy

Nine evaluations from three independent clinical cohorts had complete 2 × 2 data and a microbiological or composite reference standard applied to all participants (Table 10). Two studies with sensitivity and specificity from clinical settings were excluded from the primary pool because of partial verification. In Marquez 2025, only CAD-positive or symptom-positive participants received molecular testing. In Liao 2022, sputum testing was offered only to participants with symptoms, an abnormal CXR, known active TB or a positive interferon-γ release assay (8 TB cases: TP 8, FN 0, FP 110, TN 2,425). Partial verification cannot be corrected by back-calculation of a 2 × 2 table. Both are retained in the narrative synthesis, and Liao 2022 is analyzed as a sensitivity analysis.

#### 4.9.1 Primary pooled estimates

The pooled sensitivity was 0.896 (95% CI 0.874–0.915) and the pooled false-positive rate 0.311 (0.268–0.358), corresponding to a pooled specificity of 0.689 (95% CI 0.642–0.732) (Fig 5). The SROC AUC was 0.882 (normalized partial AUC over the observed FPR range 0.889; Fig 7). The between-evaluation standard deviations were 0.31 for logit sensitivity and 0.32 for logit FPR (correlation 0.30). The Zhou–Dendukuri *I*^2^ was approximately 75%; Holling’s sample-size-unadjusted *I*^2^ ranged from 66.2% to 88.6%, and the adjusted versions were 0.3%, which illustrates that *I*^2^ is framework-dependent in DTA meta-analysis.The 95% prediction intervals were 0.798–0.950 for sensitivity and 0.499–0.831 for specificity. The pooled sensitivity is close to the WHO 90% triage target, and its confidence interval includes it. The pooled specificity lies just below the 70% target, but its confidence interval includes 70%, and the prediction interval shows that an AI-CXR tool deployed in a new setting could plausibly have a specificity anywhere from about 50% to 83%. With only three independent cohorts, the between-cohort variance is poorly estimated and the prediction interval is approximate. The pooled specificity is an average over heterogeneous settings, not a transferable performance parameter; programs should calibrate thresholds locally.

#### 4.9.2 Accounting for non-independence and a partially verified study

Restricting the analysis to one evaluation per cohort (*k* = 3: Khan qXR, Qin qXR, Wang) gave a sensitivity of 0.879 (95% CI 0.781–0.936) and a specificity of 0.752 (0.732–0.772) (AUC 0.797). The correlation between the random effects was estimated near the boundary (0.97) and the software warned of very few primary studies, so this model is descriptive; its narrow specificity interval should not be read as precision. When Liao 2022 was added back (10 evaluations, 4 cohorts), pooled sensitivity was 0.893 (0.869–0.913), pooled specificity 0.735 (0.632–0.818), AUC 0.905 and the Zhou–Dendukuri *I*^2^ 82% (Fig 6; Table 11).

Three conclusions follow. First, pooled sensitivity changes little across these three analyses (0.879–0.896). Second, pooled specificity depends on the weighting and on one small, partially verified study: it is 0.689 when the five Dhaka algorithms (specificity 61– 74%) carry most of the weight, 0.752 when each cohort receives equal weight, and 0.735 when Liao 2022 is added back. Third, the narrow confidence intervals of the primary analysis are partly an artifact of treating algorithms evaluated on the same participants as independent; with only three independent cohorts, the uncertainty around specificity is substantial.

#### 4.9.3 Diagnostic odds ratio

The common-effect (Mantel–Haenszel) DOR was 19.11 (95% CI 18.20–20.07) and the random-effects DOR 19.20 (14.72–25.03), with substantial heterogeneity (*I*^2^ = 93.0%, τ^2^ = 0.146). These univariate summaries ignore the threshold-dependent correlation between sensitivity and specificity and are reported only for comparison with earlier reviews.

#### 4.9.4 Summary ROC curve

The SROC curve (Fig 7) shows the summary point (sensitivity 0.896, false-positive rate 0.311) within a compact 95% confidence region, and the curve extends along the FPR axis from about 0.23 to 0.42, the observed range.

### 4.10 Subgroup and sensitivity analyses

Table 11 presents the composition and results of every quantitative subgroup and sensitivity analysis. The planned analyses excluding high-risk-of-bias evaluations or data-derived thresholds were not performed: after exclusion fewer than three independent cohorts remained, and a bivariate model could not be estimated.

#### 4.10.1 Commercial versus custom models

The commercial CAD subgroup (eight evaluations from two cohorts) had a pooled sensitivity of 0.902 (0.898–0.907) and a specificity of 0.677 (0.631–0.721). Custom and other non-commercial models could not be pooled because only one (Wang 2024) had 2 × 2 data from a clinical cohort; the seven custom models reporting specificity ranged from 0.63 to 0.995, mostly on benchmark data. A pooled estimate and a descriptive range are not comparable statistical quantities, and no comparison of performance between commercial and custom models can be made from these data.

#### 4.10.2 Reference standard

None of the pooled evaluations used dataset labels, so the evidence on clinical accuracy rests entirely on culture, Xpert or composite reference standards. Culture-referenced (one cohort) and Xpert-referenced (one cohort) estimates were similar (sensitivity 0.916 and 0.902; specificity 0.677 and 0.678), but each stratum represents a single cohort and cannot separate the effect of the reference standard from that of the population. The thirteen dataset-label studies, which reported the highest accuracies, contribute only to the narrative synthesis, and their results should be regarded as technical feasibility rather than diagnostic accuracy for clinical TB.

#### 4.10.3 Dataset source and population

Public-dataset studies could not be pooled because none reported 2 × 2 data from a clinical cohort, and only one study was pediatric. These comparisons are therefore descriptive. The only pediatric model had an internal AUC of 0.903, similar to adult studies, which fell to 0.682 on external low-burden data [8].

#### 4.10.4 Internal versus external validation

In the three studies reporting both internal and external performance, AUC was consistently lower on external data: 0.878 vs 0.806 [25], 0.903 vs 0.682 [8] and 0.850 vs 0.705 [17]. The mean reduction was 0.15 AUC points (range 0.07–0.22). No formal test was performed.

### 4.11 Small-study effects

Deeks’ funnel-plot asymmetry test could not be estimated: it requires at least ten evaluations, and the primary pool has nine, drawn from three cohorts. Small-study effects were therefore not formally assessed. Selective reporting of the best-performing architecture or threshold in model-development studies remains likely.

### 4.12 Evidence for clinical translation

Each study was classified with three explicit binary criteria: (i) prospective design (consecutive or random prospective enrollment, the index test applied as prespecified and the reference standard applied to all participants); (ii) external validation (index test evaluated on data independent of model development, i.e. from a different site or population; independent evaluations of commercial software qualify); and (iii) WHO TPP performance (sensitivity ≥90% and specificity ≥70% in at least one evaluation at a reported operating point). The evidence tier was assigned mechanically, without weighting: tier A, all three criteria; tier B, two; tier C, one; tier D, none. The criteria were coded from the extraction form by one reviewer (SAM).

Three studies enrolled participants prospectively (Khan 2020, Tavaziva 2022, Liao 2022), but only the two that applied the reference standard to all participants (Khan 2020 and Tavaziva 2022, the same Karachi cohort) met criterion (i). Eight studies involved external validation, and six met the TPP in at least one evaluation (three only on internal benchmark data). Only one study (4.8%) met all three criteria: Khan 2020 (qXR). Tavaziva 2022 met two criteria but not the TPP (88.1%/57.9%); Liao 2022 reached 100% sensitivity and 95.7% specificity in active screening, but on eight TB cases and with partial verification; Marquez 2025 was a retrospective analysis of program data with partial verification; and Qin 2021 met the TPP for qXR and CAD4TB but was retrospective and used a data-derived operating point (Table 12). No study evaluated cost-effectiveness or workflow integration.

**Table 12.**
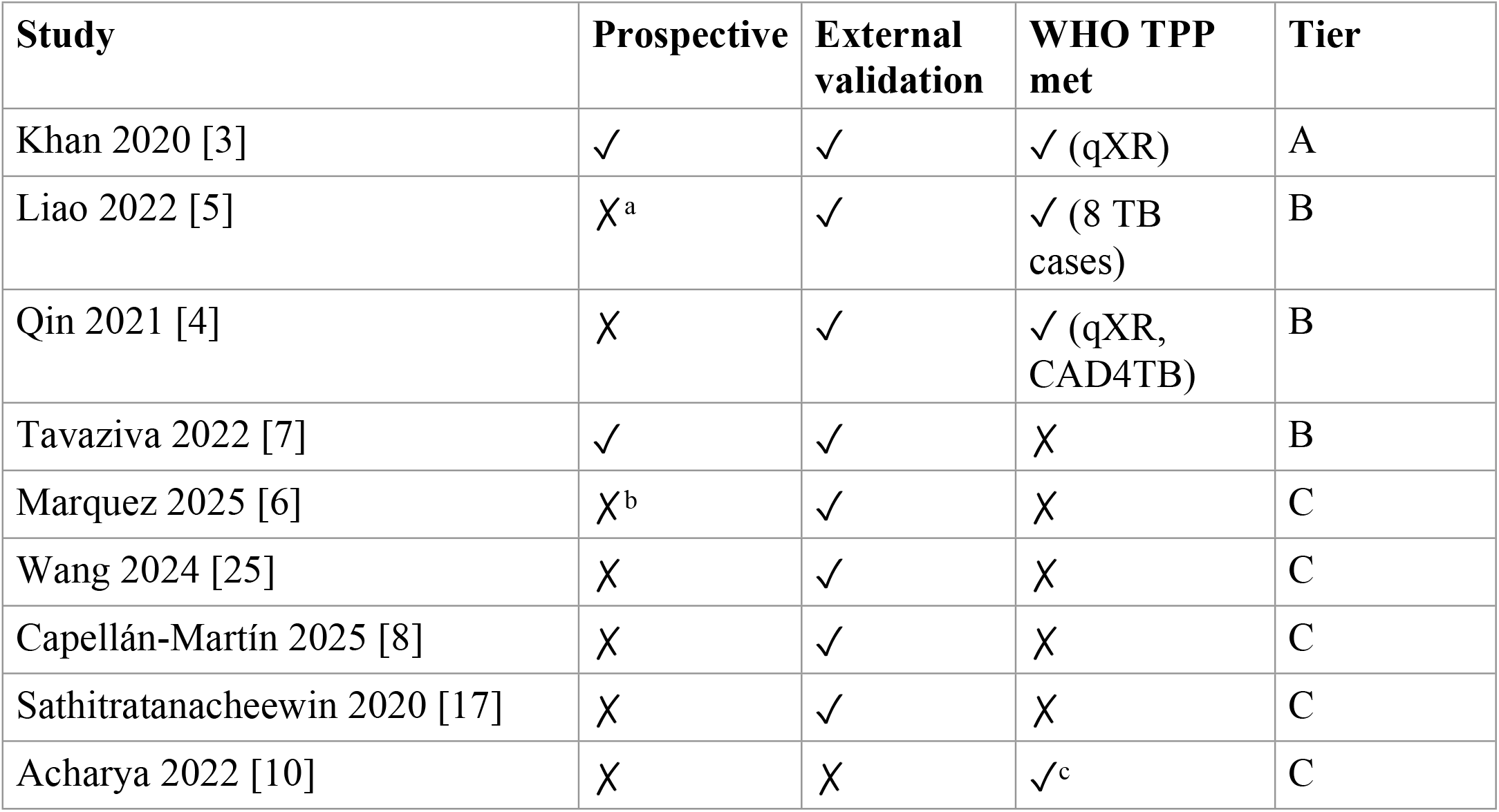
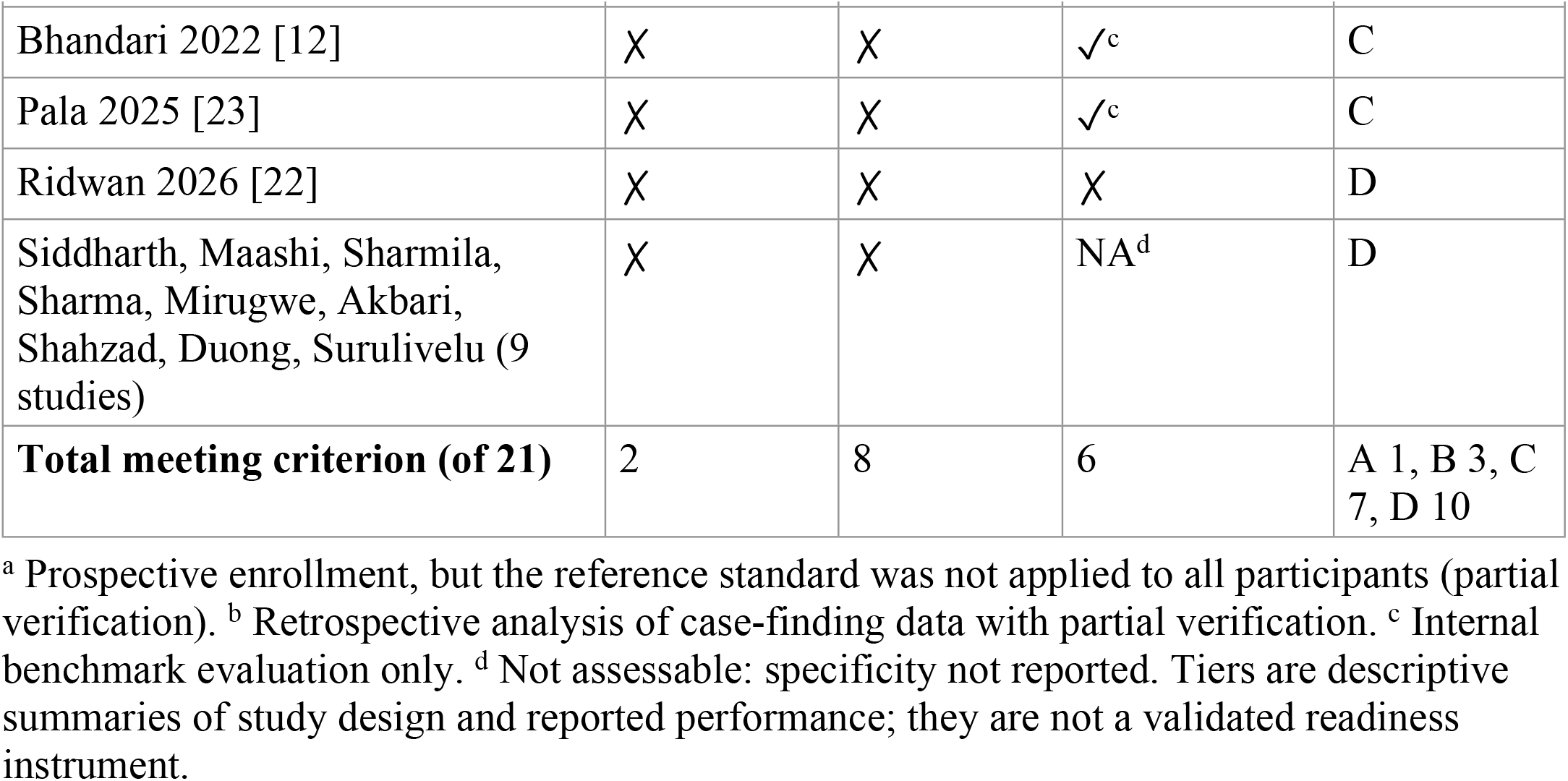
Evidence for clinical translation by study (rule-based evidence tier.

### 4.13 Certainty of evidence

Using the GRADE approach, certainty was low for the pooled sensitivity and very low for the pooled specificity (S2 File). Both were downgraded for risk of bias (seven of the nine pooled evaluations were at high overall risk) and for imprecision (each confidence interval includes its WHO threshold and only three cohorts contribute); specificity was also downgraded for inconsistency between settings.

## 5 Discussion

### 5.1 Principal findings

This review identified 21 studies (26 AI model evaluations) of AI-based pulmonary TB detection from CXR. Three findings stand out. First, in the only evaluations that could be pooled (nine evaluations of clinical cohorts with microbiological or composite reference standards applied to all participants), AI-CXR achieved a sensitivity close to the WHO triage threshold (0.896; 95% CI 0.874–0.915, which includes 0.90), with a prediction interval of 0.798–0.950. Sensitivity was broadly stable across the dependence-adjusted analyses (0.879 with one evaluation per cohort; 0.893 with Liao 2022 added back). Second, pooled specificity was 0.689, just below the 70% target; its confidence interval (0.642–0.732) includes the threshold, and its prediction interval (0.499–0.831) spans it widely. The estimate depended on weighting and on one small, partially verified study (0.752 with one evaluation per cohort; 0.735 with Liao 2022 added back). Specificity is therefore a property of the deployment setting (case mix, prior TB, age, image acquisition and operating threshold) as much as of the software. Third, the evidence base is much thinner than the number of included studies suggests. The nine pooled evaluations come from three independent cohorts; 75% of assessed evaluations were at high overall risk of bias; the thirteen dataset-label studies that report the highest accuracies do not measure accuracy for clinical TB; and only one study was prospective with complete verification, externally validated and met the TPP in at least one evaluation.

Because eight of the nine pooled evaluations share participants and only three cohorts are independent, the pooled estimates and prediction intervals are approximate summaries of a small evidence base, not performance to expect in a new hospital or program. Taken together, the data support AI-CXR as a candidate triage test whose sensitivity is reasonably consistent across clinical cohorts, not as a diagnostic replacement for microbiological testing and not as a tool with a transferable specificity. This is consistent with the use of CAD in the clinical cohorts, where AI output was used to decide who should proceed to confirmatory testing [3–6], and with WHO guidance that positions CAD as a screening and triage tool followed by a WHO-recommended rapid diagnostic test [2].

### 5.2 What the pooled estimates mean in practice

Because predictive values and referral burden depend on prevalence, it is useful to translate the pooled estimates into a population. For illustration only, in 1,000 people screened at 5% prevalence, the pooled estimates imply about 45 true positives, 5 missed cases, 295 false-positive referrals and 655 true negatives (PPV about 13%, NPV about 99%). Across the specificity prediction interval, however, false-positive referrals could range from about 160 to about 476 per 1,000 screened, an approximately three-fold difference in confirmatory-testing workload. For a program that has not calibrated its own threshold, this range, rather than the point estimate, is the relevant planning figure. It explains why the same class of software can appear efficient in one setting [4, 5] and generate large volumes of referrals in another [6], and it is why we recommend local threshold calibration against a microbiological reference standard before deployment.

### 5.3 Comparison with previous reviews

Earlier reviews of AI-based CXR analysis for TB found that most studies were development studies with case–control sampling, dataset-label reference standards and a high risk of bias [31], and more recent meta-analyses of commercial or certified products reported encouraging pooled accuracy with substantial heterogeneity [32, 33, 35]; a systematic review of machine-and deep-learning models likewise found high accuracy on benchmark data and weak reference standards [34]. Our findings agree with these reviews in two respects: sensitivity close to the triage target and specificity that varies by setting. They differ in three. First, we pooled only evaluations whose reference standard and sampling match the intended use, excluded studies with partial verification from the primary pool, and analyzed benchmark studies narratively. Second, we explicitly modeled the dependence created by evaluating several algorithms on the same participants: eight of our nine pooled evaluations shared participants, and the effective number of independent cohorts was three. Reviews that treat each algorithm as an independent study will report confidence intervals that are too narrow and, when one large head-to-head cohort contributes many algorithms, a pooled specificity that largely reflects that cohort. Third, we report prediction intervals, which show that the clinically relevant uncertainty is larger than the confidence intervals imply.

### 5.4 Model families

Commercial CAD systems provided almost all of the clinically interpretable evidence, because they were evaluated in clinical or program cohorts [3–7]. Within the Dhaka cohort, where five products read the same images, qXR v3 and CAD4TB v7 retained higher specificity than Lunit INSIGHT, InferRead DR and JF CXR-1 at the same 90% sensitivity operating point [4]; within the Karachi cohort, qXR v2 had higher specificity than CAD4TB v6 at matched sensitivity [3]. These within-cohort contrasts are the only fair comparisons available, because they hold population and reference standard constant; they concern product versions that have since been superseded and should not be generalized to current versions.

Custom CNN, transformer, hybrid and graph-based models often reported accuracy or AUC above 0.95, but almost exclusively on public benchmark datasets with balanced or multi-source case–control sampling and dataset labels [10, 12, 14, 16, 18, 20, 23, 24]. These results demonstrate technical feasibility. They do not show that custom models are more accurate than commercial CAD, because the two groups were evaluated on different populations against different reference standards; a pooled commercial estimate and a descriptive custom-model range are not comparable quantities, and we make no claim of superiority either way. Where custom models were evaluated on clinical or external data, performance was similar to or lower than that of commercial systems [8, 17, 25].

Architecture therefore explained less of the observed variation than study design, reference standard, population spectrum and validation strategy.

### 5.5 Generalizability and external validation

The most consistent finding across the narrative synthesis is a generalization gap. In the three studies reporting both internal and external performance, AUC fell by a mean of 0.15 (range 0.07–0.22) on external data [8, 17, 25]; the pediatric model fell from 0.903 to 0.682 in a low-burden external cohort [8]; and a model trained on Shenzhen data lost substantial performance on an unrelated external source [17]. Commercial software is not exempt: the same product family that met the TPP in Karachi and in Dhaka had a specificity of 28.1% at a fixed threshold in Philippine case finding [3, 4, 6], although partial verification in that study limits interpretation. Performance on a public benchmark should be reported as technical feasibility; claims of clinical accuracy require external validation on independent, consecutively sampled clinical data [48–50].

### 5.6 Dataset quality and reference standards

The highest reported accuracies came from datasets with the weakest link to clinical TB. Public compilations frequently mix sources (for example, TB images from one country and normal images from another), which allows models to learn acquisition-related shortcuts rather than disease; several compilations re-use the same Shenzhen, Montgomery, Belarus and NIAID images, so “different” benchmarks are not independent; and labels are often radiographic or of unknown provenance. Culture or a well-defined composite reference standard was used in only five studies, all of them clinical cohorts [3, 5, 7, 8, 25]. A radiographic label measures agreement with a reader, not detection of TB, and so such studies answer a different question from the one relevant to screening programs. Future datasets should include consecutive or random sampling, demographics, HIV status, prior TB, smear and culture status, projection and scanner details, and complete reference-standard data.

### 5.7 Explainability

Five studies used XAI methods (Table 9), but only one compared explanations with expert radiologists [12], and none tested whether explanations changed clinical decisions or helped users detect model errors. Saliency maps are known to be unstable and can highlight plausible regions even when the model relies on shortcuts. Explainability should therefore be treated as one component of a broader validation framework (calibration, subgroup performance, robustness to acquisition differences and post-deployment monitoring), not as evidence of clinical reasoning.

### 5.8 Clinical and public-health implications

For programs, the evidence supports three practical conclusions. First, AI-CXR can be considered as a triage step before WHO-recommended rapid diagnostic testing in adults, where its sensitivity is reasonably consistent across clinical cohorts [2]. Second, because specificity and hence referral burden are setting-specific, thresholds should be calibrated locally on a sample with microbiological reference testing, with explicit targets for the acceptable miss rate and confirmatory-testing capacity [4, 6]. Third, subgroups in which performance may differ (children, people living with HIV, people with prior TB, older adults and smear-negative disease) remain poorly characterized: the Karachi cohort was 99.5% HIV-negative, no study reported HIV-stratified accuracy, and only one study was pediatric [3, 7, 8]. A single universal threshold is unlikely to perform equally well in these groups.

### 5.9 Reporting standards

The most frequently missing items were complete 2 × 2 tables (available for only 11 of 26 evaluations), specificity (reported for 17 of 26), predictive values, calibration, prespecified threshold rules, subgroup results, dataset provenance and code availability. Adherence to STARD 2015, STARD-AI and TRIPOD+AI [48–50] would allow future evaluations to be pooled; in particular, reporting 2 × 2 counts at prespecified thresholds and the number of participants per cohort would allow dependence between evaluations to be modeled properly.

### 5.10 Strengths and limitations

Strengths of this review include a transparent account of every evaluation across all analyses (S1 Appendix, Table B), restriction of pooling to evaluations matching the intended use and with complete verification, explicit handling of non-independence, prediction intervals, and a sensitivity analysis showing how one partially verified study changes the pooled specificity.

The review also has important limitations. (i) *Search.* Only English-language publications were included, which may omit evidence from high-burden settings publishing in other languages. Five sources were searched on 15 March 2026, so later publications are not captured. Scopus, Web of Science, Embase, Google Scholar and Dimensions were not searched; for Lens.org only the first 100 records were screened. The executed search strings did not include “computer-aided detection/diagnosis”, “automated detection”, “digital radiography” or commercial product names as separate terms, so implementation studies indexed only under CAD terminology may have been missed. A post hoc check of PubMed with the omitted terms, covering January 2020 to October 2026, found at least eleven further clinical evaluations that were not screened for this review; some were published after the original search date and would fall outside its date limits. Examples are multi-country, prospective and Xpert-or culture-referenced evaluations of commercial CAD [51–56], two case–control evaluations [57, 58], studies with partial verification [59, 60] and an independent evaluation of 12 commercial products against Xpert MTB/RIF [61].

Consequently the review is incomplete for clinical-cohort evaluations of commercial CAD, and the pooled estimates and the number of independent cohorts would probably change if these were added. Updating the search and re-estimating the pooled results is the most important next step. (ii) *Protocol.* No protocol was prepared or registered, and several analyses were added post hoc or in response to peer review (Table 1); the risk of selective analysis cannot be excluded, although all such analyses are reported in full. (iii) *Single-reviewer conduct.* Screening, full-text assessment, data extraction and risk-of-bias assessment were performed by one reviewer; a co-author also reviewed the work informally, but the check was not documented and no independent duplicate assessment or agreement statistic is available, which increases the risk of selection and extraction errors. Reasons for full-text exclusion were recorded for only 14 of 31 reports, and the duplicate count (44) was not independently verified. (iv) *Few independent cohorts.* The meta-analysis rests on three independent cohorts, which limits precision, makes the between-cohort variance poorly estimated, prevents meaningful meta-regression and rules out a multilevel model; the prediction intervals are approximate. (v) *Dependence.* The within-cohort sampling covariance between algorithms read on the same images could not be estimated from published data. (vi) *Operating points.* Several evaluations used thresholds chosen to achieve 90% sensitivity in the test data, which inflates sensitivity and makes pooled sensitivity partly a design choice; an analysis excluding data-derived thresholds could not be estimated. (vii) *Small-study effects*.

Deeks’ test requires ten or more evaluations and could not be estimated, and selective reporting of the best architecture or threshold is likely in development studies. (viii) *Aggregate data.* Study-level data precluded assessment of patient-level effect modification by age, sex, HIV status, smear status, prior TB or image projection. (ix) *Quality ratings*.

QUADAS-2 judgments involve subjectivity, and two studies could not be assessed. (x) *Partial verification.* Excluding Liao 2022 and Marquez 2025 removes two screening settings from the pool; this was judged necessary, but it narrows the evidence on community screening. Finally, commercial software versions evolve rapidly, so estimates for specific products may not apply to current versions.

## 6 Future work

*Prospective multicenter evaluation.* Future studies should enroll consecutive or random samples of the intended screening or triage population, apply a microbiological reference standard to all participants (or correct for partial verification), and report sensitivity, specificity, predictive values and downstream testing burden at prespecified thresholds [2, 48].

*External validation.* Claims of clinical usefulness should require evaluation on geographically and technically distinct data, including different prevalence settings, scanners and projections (PA, AP and portable AP), since projection affected performance in the emergency-department study [25].

*Standardized reporting.* Primary studies should report complete 2 × 2 tables with confidence intervals, ROC and precision–recall curves, calibration, threshold selection rules, subgroup analyses, dataset provenance and code, in accordance with STARD-AI and TRIPOD+AI [49, 50].

*Pediatric and HIV populations.* Larger and more diverse pediatric studies, and studies reporting accuracy by HIV status (with CD4 count where available), smear status and prior TB, are needed [8].

*Validated explainability.* XAI outputs should be evaluated against radiologist-defined abnormalities with inter-reader agreement, compared between correct and incorrect predictions, and tested for their effect on clinical decisions.

*Economic and implementation evaluation.* Cost-effectiveness analyses should compare AI-CXR pathways with human reading, symptom screening and universal molecular testing, and report cost per case detected, confirmatory tests avoided, missed cases and time to diagnosis. Implementation studies should assess workflow integration, acceptability, quality assurance, model drift and linkage to treatment, with periodic local re-validation after deployment.

## 7 Conclusion

AI-based interpretation of chest radiographs shows promising diagnostic performance for pulmonary TB triage in clinical cohorts. Pooled sensitivity was 0.896 (95% CI 0.874–0.915), close to the WHO 90% target, and was stable across the dependence-adjusted analyses. Pooled specificity was 0.689 (95% CI 0.642–0.732); its point estimate is just below the WHO 70% target, but its confidence interval includes 70%. The data therefore show neither that these targets are met nor that they are missed. Specificity varied with the product, the setting and the studies included, so the pooled value should not be used as a benchmark or transferred to new settings. The pooled evidence rests on only three independent cohorts, 75% of assessed evaluations were at high risk of bias, certainty of evidence was low for sensitivity and very low for specificity, the most impressive accuracies came from benchmark datasets with dataset-label reference standards, and the search missed eligible clinical studies. Only 1 of 21 studies was prospective with complete verification, externally validated and met the WHO thresholds. Current evidence does not demonstrate clinical maturity for widespread deployment, and the pooled estimates are hypothesis-generating. AI-CXR tools should be deployed, if at all, as triage aids accountable to microbiological confirmation, with local threshold calibration and ongoing monitoring, while prospective, externally validated evaluations are conducted.

## Data Availability

We have shared.

## Acknowledgments

The first author ran all analyses and the corresponding author checked every output and takes responsibility for the content.

## Supporting information

**S1 Checklist. PRISMA 2020 checklist.**

**S2 Checklist. PRISMA-DTA checklist.**

**S1 Appendix. Search strategies, data extraction form, QUADAS-2 signalling questions with per-evaluation judgements (Fig A), and master accounting table (Table B).**

**S1 File. Extraction data, R analysis script and README (ZIP archive).**

**S2 File. Certainty of evidence (GRADE) table.**

## Notes

### Competing Interest Statement

The authors have declared no competing interest.

